# Bridging the gap: a prospective trial comparing programmable targeted long-read sequencing and short-read genome sequencing for genetic diagnosis of cerebellar ataxia

**DOI:** 10.1101/2024.07.08.24309939

**Authors:** Haloom Rafehi, Liam G. Fearnley, Justin Read, Penny Snell, Kayli C. Davies, Liam Scott, Greta Gillies, Genevieve C. Thompson, Tess A. Field, Aleena Eldo, Simon Bodek, Ernest Butler, Luke Chen, John Drago, Himanshu Goel, Anna Hackett, G. Michael Halmagyi, Andrew Hannaford, Katya Kotschet, Kishore R. Kumar, Smitha Kumble, Matthew Lee-Archer, Abhishek Malhotra, Mark Paine, Michael Poon, Kate Pope, Katrina Reardon, Steven Ring, Anne Ronan, Matthew Silsby, Renee Smyth, Chloe Stutterd, Mathew Wallis, John Waterston, Thomas Wellings, Kirsty West, Christine Wools, Kathy H. C. Wu, David J. Szmulewicz, Martin B. Delatycki, Melanie Bahlo, Paul J. Lockhart

**Affiliations:** Population Health and Immunity Division, The Walter and Eliza Hall Institute of Medical Research, Royal Pde, Parkville, Victoria 3052, Australia; Department of Medical Biology, University of Melbourne, Royal Pde, Parkville, Victoria 3052, Australia; Bruce Lefroy Centre, Murdoch Children’s Research Institute, Royal Children’s Hospital, Flemington Rd, Parkville, Victoria 3052, Australia; Department of Neuroscience, Central Clinical School, Monash University, The Alfred Centre, Commercial Rd, Melbourne, Victoria 3004, Australia; Department of Paediatrics, University of Melbourne, Royal Children’s Hospital, Flemington Rd, Parkville, Victoria 3052, Australia; Austin Health, Studley Rd, Heidelberg, Victoria 3084, Australia; Peninsula Health, Hastings Rd, Frankston, VIC 3199, Australia; Department of Neurology, Alfred Hospital, Commercial Rd, Melbourne, Victoria 3004, Australia; Department of Medicine, St Vincent’s Hospital, University of Melbourne, Victoria Pde, Fitzroy, Victoria 3065, Australia; Florey Institute of Neuroscience and Mental Health, Royal Pde, Parkville, Victoria 3052, Australia; Hunter Genetics, Hunter New England Health Service, Turton Rd & Tinonee Rd, Waratah, New South Wales 2298, Australia; University of Newcastle, University Dr, Callaghan, New South Wales 2308, Australia; Neurology Department, Royal Prince Alfred Hospital, Missenden Rd, Camperdown, New South Wales 2050, Australia; Central Clinical School, University of Sydney, Johns Hopkins Dr, Camperdown, New South Wales 2050, Australia; Department of Neurology, Westmead Hospital, Hawkesbury Rd & Darcy Rd, Westmead, New South Wales 2145, Australia; Brain and Nerve Research Centre, Concord Clinical School, University of Sydney, Mallett St, Camperdown, New South Wales 2050, Australia; Department of Neurology, Concord Repatriation General Hospital, Hospital Rd, Concord, New South Wales 2139, Australia; Department of Clinical Neurosciences, St Vincent’s Hospital, Victoria Pde, Fitzroy, Victoria 3065, Australia; Molecular Medicine Laboratory and Neurology Department, Concord Repatriation General Hospital, Hospital Rd, Concord, New South Wales 2139, Australia; Faculty of Medicine and Health, The University of Sydney, Science Rd, Camperdown, New South Wales 2050, Australia; Genomics and Inherited Disease Program, The Garvan Institute of Medical Research, Victoria St, Darlinghurst, New South Wales 2010, Australia; School of Medicine, University of New South Wales, High St & Botany St, Sydney, New South Wales 2052, Australia; Victorian Clinical Genetics Services, Murdoch Children’s Research Institute, Royal Children’s Hospital, Flemington Rd, Parkville, Victoria 3052, Australia; Department of Clinical Genetics, Austin Health, Studley Rd, Viewbank, Victoria 3084, Australia; Department of Neurology, Launceston General Hospital, Charles St, Launceston, Tasmania 7250, Australia; Department of Neuroscience, University Hospital Geelong, Bellerine St, Geelong, Victoria 3220, Australia; Department of Neurology, Royal Brisbane and Women’s Hospital, Butterfield St, Herston, Queensland 4006, Australia; Neurology Footscray, Eleanor St, Footscray, Victoria 3011, Australia; Department of Neurology, St Vincent’s Hospital Melbourne, Victoria Pde, Victoria 2065, Australia; Albury Wodonga Specialist Centre, Ramsay Pl, West Albury, New South Wales 2640, Australia; Newcastle Medical Genetics, Grainger St, Lambton, New South Wales 2299, Australia; St Vincent’s Clinical Genomics, St Vincent’s Hospital, Boundary St, Darlinghurst, New South Wales 2010, Australia; Tasmanian Clinical Genetics Service, Tasmanian Health Service, Royal Hobart Hospital, Liverpool St, Hobart, Tasmania 7001, Australia; School of Medicine and Menzies Institute for Medical Research, University of Tasmania, Liverpool St, Hobart, Tasmania 7000, Australia; Department of Neurology, John Hunter Hospital, Lookout Rd, New Lambton Heights, New South Wales 2305, Australia; Genomic Medicine, The Royal Melbourne Hospital, Grattan St, Parkville, Victoria 3052, Australia; Department of Neurology, Calvary Health Care Bethlehem, Kooyong Rd, Caulfield South Victoria 3162, Australia; Department of Neurology, The Royal Melbourne Hospital, Royal Pde, Parkville, Victoria 3052, Australia; School of Medicine, University of Notre Dame, Oxford St, Darlinghurst, New South Wales 2010, Australia; Discipline of Genomic Medicine, Faculty of Medicine and Health, University of Sydney, Science Rd, Camperdown, New South Wales 2050, Australia; Royal Victorian Eye and Ear Hospital, Gisborne St, East Melbourne, Victoria 3002, Australia; Bionics Institute, Albert St, East Melbourne, Victoria 3002, Australia

**Keywords:** cerebellar ataxia, repeat expansion, short tandem repeats, long-read sequencing, short-read genome sequencing

## Abstract

The cerebellar ataxias (CA) are a heterogeneous group of disorders characterized by progressive incoordination. Seventeen repeat expansion (RE) loci have been identified as the primary genetic cause and account for >80% of genetic diagnoses. Despite this, diagnostic testing is limited and inefficient, often utilizing single gene assays. This study evaluated the effectiveness of long- and short-read sequencing as diagnostic tools for CA. We recruited 110 individuals (48 females, 62 males) with a clinical diagnosis of CA. Short-read genome sequencing (SR-GS) was performed to identify pathogenic RE and also non-RE variants in 356 genes associated with CA. Independently, long-read sequencing with adaptive sampling (LR-AS) and performed to identify pathogenic RE. SR-GS identified pathogenic variants in 38% of the cohort (40/110). RE caused disease in 33 individuals, with the most common condition being SCA27B (n=24). In comparison, LR-AS identified pathogenic RE in 29 individuals. RE identification for the two methods was concordant apart from four SCA27B cases not detected by LR-AS due to low read depth. For both technologies manual review of the RE alignment enhanced diagnostic outcomes. Orthogonal testing for SCA27B revealed a 16% and 0% false positive rate for SR-GS and LR-AS respectively. In conclusion, both technologies are powerful screening tools for CA. SR-GS is a mature technology currently utilized by diagnostic providers, requiring only minor changes in bioinformatic workflows to enable CA diagnostics. LR-AS offers considerable advantages in the context of RE detection and characterization but requires optimization prior to clinical implementation.

## INTRODUCTION

Genetic technologies are transforming healthcare by enabling genomic medicine, an emerging discipline that utilizes genetic data to improve clinical care and outcomes. Next Generation Sequencing (NGS) is an important tool for genomic medicine, underpinning discovery, diagnostics, and understanding of disease mechanisms (Rehm 2017). Genomic medicine provides diagnostic certainty, key information for prognosis, genetic counseling, reproductive planning and facilitates the development and delivery of improved treatments targeted to disease mechanisms (McCarthy et al. 2013; Garcia-Foncillas et al. 2021). There is a significant treatment pipeline for personalized therapies, including gene and pharmacological therapies. For instance, treatment for spinocerebellar ataxia 27B (SCA27B), shows promising therapeutic outcomes with 4-aminopyridine (4-AP) (Wilke et al. 2023). However, genetic technologies have had limited success in extending genomic medicine’s benefits to disorders caused by pathogenic nucleotide repeat expansions (RE).

RE disorders typically have significant neurological and/or neuromuscular outcomes. They occur when a segment of repetitive DNA, termed a short tandem repeat (STR), expands beyond a gene-specific threshold. STR are composed of tandem arrays of 1–12 base pair sequence motifs and constitute ∼6% of the human genome (Willems et al. 2014; Mousavi et al. 2019). To date the genetic basis of 79 RE disorders have been described (reviewed in (Cortese et al. 2023; Rajan-Babu et al. 2024)), including polyglutamine disorders such as Huntington disease, fragile X syndrome and hereditary cerebellar ataxias. RE can occur within exonic, intronic, untranslated or intergenic regions and are associated with various pathogenic mechanisms including loss-of-function, gain-of-function, transcriptional dysregulation, protein misfolding/aggregation and repeat-associated non-AUG (RAN) translation. Some disorders result from combinations of these mechanisms (reviewed in (Depienne and Mandel 2021)). Collectively, repeat expansion disorders cause some of the most common genetic disorders seen by neurologists (Paulson 2018). Moreover, the RE-mediated disease burden is significantly underestimated. RE are difficult to amplify using standard molecular technologies such as PCR (Schlotterer and Tautz 1992) and there is evidence suggesting that additional RE remain to be identified (Depienne and Mandel 2021). In addition, RE are often located in non-coding DNA, ‘hidden’ from gene, panel- and exome-based discovery approaches. Pathogenic RE embedded deep within nonpathogenic STR (Rosenbohm et al. 2022) or composed of novel motifs present additional challenges (Dolzhenko et al. 2020). The timeline of RE identification demonstrates the issues. While the first pathogenic RE were described in 1991 (La Spada et al. 1991; Oberle et al. 1991; Verkerk et al. 1991), approximately 50% of pathogenic RE identified to date have been described in the last ten years. The acceleration in discovery has been driven largely by the development of PCR-free short-read and long-read genomic sequencing technologies and associated bioinformatic tools (Depienne and Mandel 2021; Gall-Duncan et al. 2022; Read et al. 2023).

RE disorders also challenge diagnostic service providers, impacting the implementation of genomic medicine. One issue is the relatively low incidence of some RE disorders; in many settings it is not economically viable to provide current, RE-specific diagnostic testing for loci/conditions with low prevalence (Stevanovski et al. 2022). In addition, technological challenges exist in the molecular characterization of expanded repeat DNA. Traditional diagnostic techniques such as Southern blot, PCR sizing and repeat-primed PCR analysis (RP-PCR) are labor-intensive, imprecise and do not scale easily to large cohorts requiring testing; generally, each individual RE requires a separate assay with specific probes or primers (Bahlo et al. 2018). Exome analysis using short-read next generation sequencing (NGS) has become a mainstay diagnostic methodology for diagnosing pathogenic single nucleotide variants (SNV), small insertions and deletions (indel) or copy number variants (CNV) but is yet to be widely implemented in RE diagnostics (Lappalainen et al. 2019). Moreover, like other high throughput diagnostic technologies such as multiplex amplicon sequencing and targeted gene panel analysis, exome sequencing utilizes PCR amplification of target sequences. While bioinformatic tools now available in the research space allay concerns about the effects of PCR stutter and non-unique mapping of repetitive NGS reads (Bahlo et al. 2018; Tankard et al. 2018), they are yet to be widely implemented in diagnostic pipelines. Another shortcoming of exome sequencing for diagnosing pathogenic RE is that most loci are outside coding regions and therefore not captured by current ‘off-the-shelf’ library preparation kits.

Hereditary cerebellar ataxias (CA) exemplify the diagnostic and healthcare challenges of RE disorders. CA are a heterogeneous group of rare, incurable and often life-limiting disorders characterized by progressive incoordination (Jayadev and Bird 2013). There are approximately 100 clinically recognized CA, with similar numbers caused by autosomal dominant and autosomal recessive mechanisms. The prevalence of these disorders varies widely depending on genetic ancestry and geographical location, with estimates of a global average of ∼6:100,000 (Ruano et al. 2014; Rudaks et al. 2024). These debilitating disorders predominantly impact locomotion, hand coordination, speech, swallowing and vision. Apart from the recent approval of omaveloxolone for Friedreich ataxia (FRDA)(Lee 2023) there are no disease-modifying treatments for these conditions. Treatment consists of symptom management, such as using adaptive devices and ongoing physical and occupational therapy (de Silva et al. 2019). The predominant genetic cause of CA, accounting for over 80% of diagnoses, is a pathogenic RE in one of the 17 loci associated with CA identified to date. Non-RE pathogenic variants in ∼100 other genes also contribute to disease prevalence (Beaudin et al. 2019; Rudaks et al. 2024), presenting a daunting panel of unusual variants and genes requiring examination. Diagnostic testing methodologies and outcomes for CA vary broadly. In Australia, standard clinical diagnostic testing utilizes single PCR/capillary array assays for between five and seven RE loci. Our centre, servicing a population of ∼6 million, only tests for six RE causing CA (spinocerebellar ataxia (SCA)1/2/3/6/7 and FRDA) with a diagnostic rate of ∼5% (unpublished data 2015-2022). A higher diagnostic rate can be achieved by utilizing multiple testing methodologies to ensure evaluation of multiple genetic variant types, including RE, SNV/indel and CNV, with yields ranging between ∼30-60% (Hadjivassiliou et al. 2017; Kang et al. 2019). However, these comprehensive, long-term studies were performed in a research setting. Clinical service providers prefer to deploy a single frontline test with high sensitivity and specificity to maximize yield and minimize cost. Notably, the yield for CA with short-read exome sequencing, arguably the most utilized and cost effective frontline diagnostic test, is only ∼25% (Rexach et al. 2019; Ngo et al. 2020).

Two technologies have recently been proposed as potential rapid and comprehensive diagnostic methods for RE disorders. Single-molecule long read sequencing using adaptive sampling (LR-AS, Oxford Nanopore Technologies) enables user-defined target selection in real time (Payne et al. 2021). This technology allows simultaneous screening of multiple target loci, capturing up to ∼4% of the genome. Two recent proof of concept pilot studies have demonstrated the method’s potential utility to diagnose RE disorders by simultaneous analysis of 37 (Stevanovski et al. 2022) or 59 (Miyatake et al. 2022a) RE loci in small retrospective cohorts of 37 and 22 patients, respectively. Alternatively, short-read genome sequencing (SR-GS) is emerging as a potential replacement for exome sequencing in rare disorders. This shift is driven by recognition of the gains achievable in diagnostic rates and the decreasing cost and availability of genome sequencing. Standard diagnostic analysis pipelines previously had limited capability to assess RE loci (Ashley 2016; Turro et al. 2020), however recent advances in bioinformatic tools and technologies mean that it is now feasible to efficiently detect both non-RE and RE pathogenic variants in SR-GS (Leitao et al. 2024). Notably, a large study recently demonstrated high sensitivity and specificity (>97%) to detect 13 pathogenic RE in a retrospective cohort of 404 patients. The prospective analysis of these 13 RE in 11,631 patients identified 81 RE with a false discovery rate of 16% (Ibanez et al. 2022).

Here, we report a prospective trial comparing diagnostic outcomes of standard clinical testing in the Australian context (five pathogenic RE) with both short-read and long-read technologies in a cohort of 110 individuals with a clinical diagnosis of cerebellar ataxia and clearly demonstrate the substantial diagnostic gains achievable for individuals and families affected by CA.

## RESULTS

### Participant recruitment and details

The study design is summarized in Figure 1. A cohort of 110 individuals with a clinical diagnosis of CA, referred for diagnostic testing, were recruited to the research program over a two year period (2022-2023). Individuals were excluded if there was clinical suspicion of an acquired cause of ataxia, based on history of acute injury or illness, toxic exposure, or rapid onset. All participants were singletons, 17% had a family history of ataxia and included 48 female/62 male individuals with adult-onset ataxia. The mean age at onset was 56±14 years (range 15-77) and mean age at testing 68±13 years (range 29-88). All individuals completed diagnostic testing for five pathogenic RE in SCA1, SCA2, SCA3, SCA6 and SCA7 prior to research based genetic testing. Any individuals with a positive diagnostic test result were excluded from the trial.

**Figure 1.**
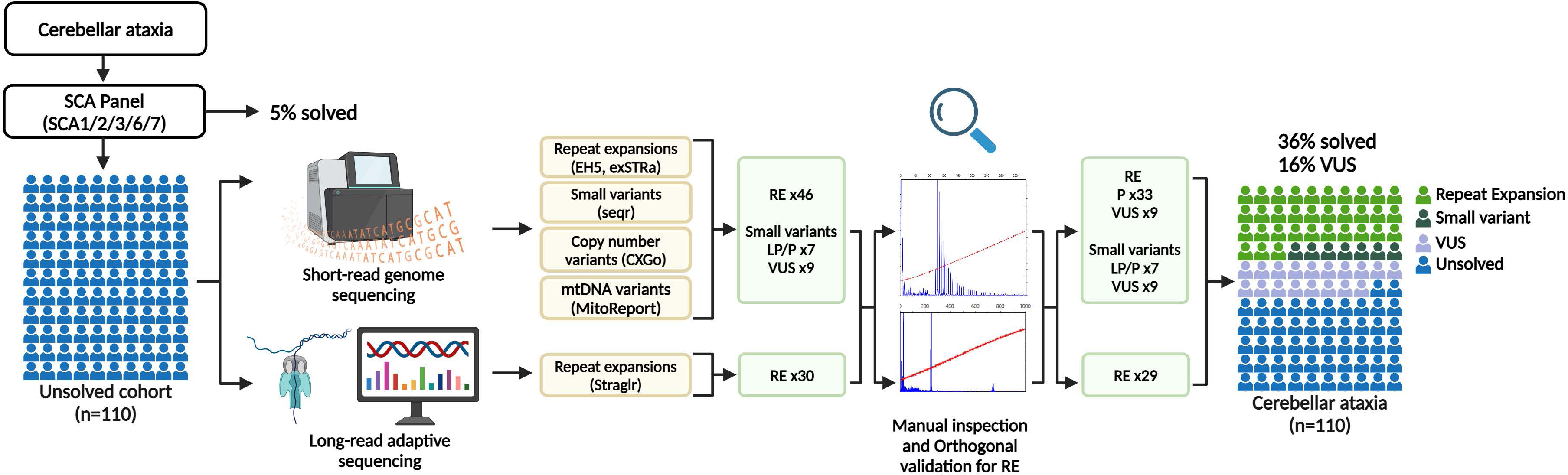
Overview of the study and investigations performed. SCA, spinocerebellar ataxia; EH5, ExpansionHunter5; RE, repeat expansion; LP, likely pathogenic; P, pathogenic; VUS, variant of uncertain significance.

### RE identification with short-read genome sequencing (SR-GS)

The primary purpose of this trial was to compare the performance of SR-GS and LR-AS for the identification of pathogenic RE. At the time of trial initiation there were 67 known pathogenic RE, 17 directly associated with CA and the remainder with a broader range of neurogenetic, neuromuscular and other health conditions. Therefore, all 67 RE were interrogated as part of this program. However, given that the clinical indication for study inclusion was CA, a specific focus of the analysis were the 22 RE listed in Table 1. This included the 17 RE associated with CA and five additional pathogenic RE that are a relatively frequent cause of disease and are also offered as a diagnostic test at our center (Victorian Clinical Genetics Services, VCGS).

**Table 1.**
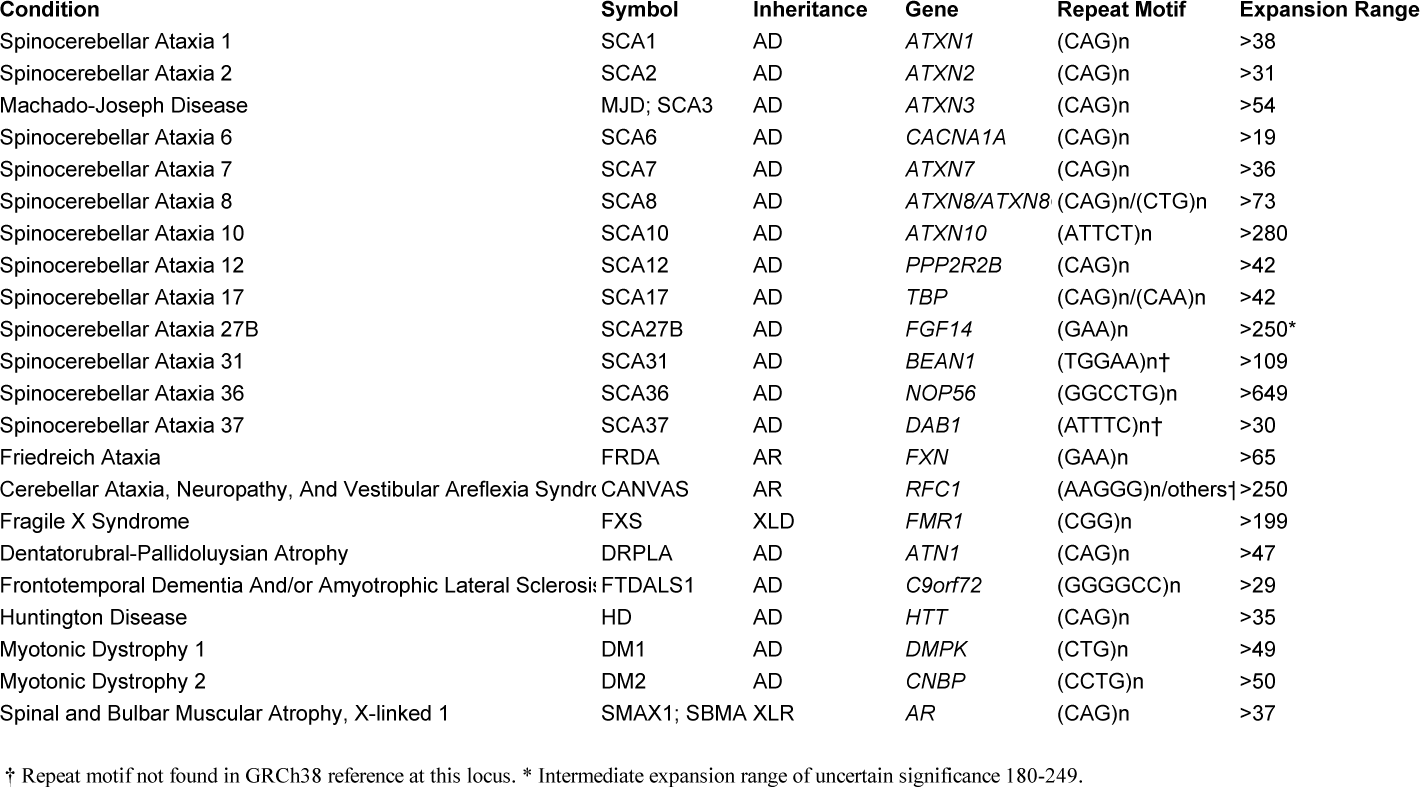
Repeat expansion target loci for cerebellar ataxias (CA) and clinically significant non-cerebellar ataxia conditions (non-CAs), adapted from (Tankard et al. 2018) and (Depienne and Mandel 2021).

ExpansionHunter5 (EH5) (Dolzhenko et al. 2019) was used to genotype the 22 RE loci in the SR-GS data. A post-analysis custom kmer filter was used to remove genotype calls where the predominant motif detected in the reads does not match the motif tested by EH5 (Fearnley et al. 2024). All individuals with potential expansions were also manually reviewed in the Integrative Genomics Viewer (IGV) to confirm motif composition. We detected two individuals with dominant *ATXN8OS* expansions greater than the pathogenic threshold for SCA8 (Figure 2A).

**Figure 2.**
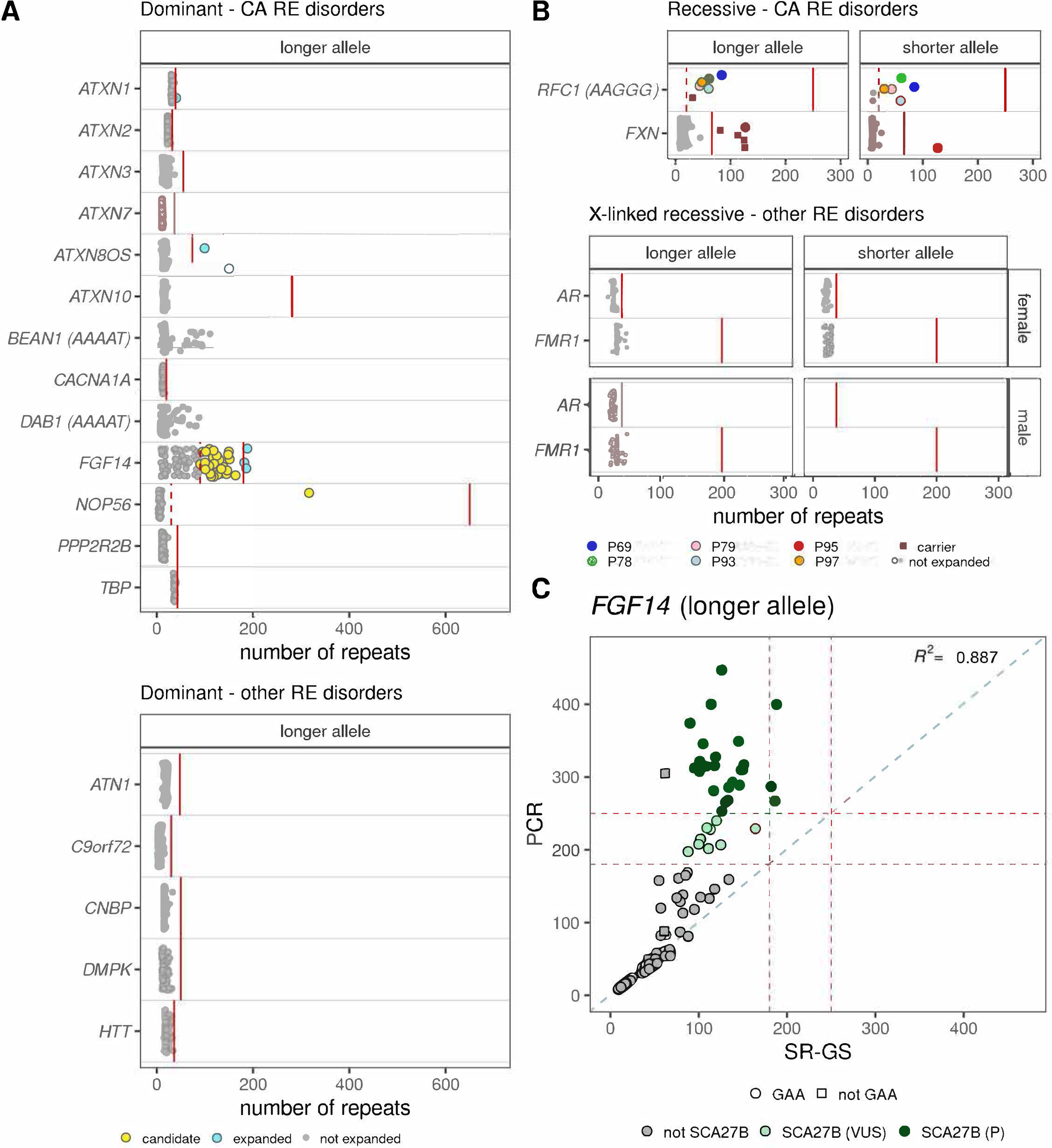
Targeted repeat expansion screening in SR-GS identifies potential RE diagnoses. STR genotypes were determined for short-listed loci with EH5. A. Genotypes are shown for the longer allele in dominant RE disorders that cause ataxia (top) and those that cause other disorders (bottom). Blue circles indicate individuals with an expansion in the respective allele that is larger than the pathogenic threshold (solid red line), while yellow circles are individuals who exceed an EH5-specific threshold (dashed red line) and are candidates for further investigation. B. Genotypes for the shorter and longer alleles are shown for autosomal recessive RE ataxia disorders (top) and X-linked recessive disorders other than ataxia, split by sex (bottom). Individuals who are heterozygous carriers for an allele expanded beyond the pathogenic threshold (solid red line) or an allele larger than the EH5-specific threshold (dashed red line) are shown as brown squares. Individuals who carry two alleles expanded beyond either the pathogenic or EH5 specific threshold are shown as coloured circles. C. Concordance plot showing a comparison of the *FGF14* STR EH5 genotypes from SR-GS compared to PCR sizing (longer allele only). The pathogenic (>250 repeats) and VUS (180 repeats) thresholds are shown as dashed red lines. A 1:1 correlation is shown as a dashed blue line. The R2 is a Pearson’s correlation. Dark green circles are individuals with a confirmed SCA27B diagnosis (P) and those in light green are SCA27B VUS. Circles indicate GAA motifs and squares are non-GAA motifs, determined by observation in IGV.

One individual was found to have an expanded allele for *NOP56*, however the expansion (317 repeats) is lower than the pathogenic threshold (650 repeats) for SCA36 (Figure 2A). We have previously shown that expansions in *NOP56* are typically underestimated by EH5 due to their large size vastly exceeding the read length (Rafehi et al. 2020). In addition, the *NOP56* STR is stable in the general population, with most alleles reported in gnomAD in the 4-10 repeats range, and only three alleles between 15-21 repeats. Therefore, any individual with a NOP56 expansion ≥30 repeats has further testing to confirm or exclude a SCA36 diagnosis. Manual review confirmed a GGGCCT expansion at the locus.

*FGF14* STR size is also known to be underestimated by RE calling in SR-GS data. Our previous work identified high concordance between PCR and SR-GS sizing for STR up to ∼(GAA)_100_, however larger RE are typically underestimated by RE genotyping tools such as EH5 (Rafehi et al. 2023). In contrast to *NOP56*, in which outliers are easy to detect, the *FGF14*-GAA STR is highly unstable and there is significant variation in the length in the general population. As a result, it is not possible to distinguish pathogenic expansions (≥250) from alleles of ∼100-249 using EH5. Based on previous experience with this locus, we prioritize any individual with a EH5 genotype call of ≥ 90 repeats as candidates for SCA27B. Using this approach, we identified 37 individuals suspected to have SCA27B (Figure 2A). Manual review confirmed that all 37 RE were composed of pure (GAA)n.

Expanded sequences were also detected for the benign reference motif (AAAAT) in *BEAN1* and *DAB1* (Figure 2A) but neither of the pathogenic motifs (TGGAA) or (ATTTC) were identified in the short-read data. Although expansions of the reference motif alone are not pathogenic for these two loci, we cannot exclude the possibility that these individuals have a pathogenic motif embedded deep within the reference motif as these cannot be detected with SR-GS (Rosenbohm et al. 2022).

Analysis of recessive loci also identified potential pathogenic RE in *FXN* and *RFC1*. One individual has two expanded alleles in *FXN*, consistent with a diagnosis of FRDA (Figure 2B). We also identified four individuals with a single expanded *FXN* allele (Supplemental_Table_S1.xls). We tested for the possibility of a second pathogenic small variant or CNV on the other *FXN* allele, however no candidate variants were identified, suggesting these individuals are heterozygous carriers. Testing for the non-reference pathogenic AAGGG motif in *RFC1* did not identify any individuals with a RE greater than the pathogenic threshold (>250 repeats (Dominik et al. 2023)). However, we have previously shown that AAGGG RE sizing of *RFC1* is underestimated in SR-GS (Rafehi et al. 2019). Therefore, any individual with an AAGGG allele ≥30 repeats is considered likely to carry a pathogenic RE in *RFC1*. Using this threshold, we identified five individuals with pathogenic biallelic RE in *RFC1*, consistent with the clinical presentation of cerebellar ataxia, neuropathy and vestibular areflexia syndrome (CANVAS) in these individuals (Table 2). In addition, we identified three individuals with a heterozygous pathogenic AAGGG *RFC1* allele (Supplemental_Table_S1.xls) but failed to identify a second pathogenic variant on the other allele, suggesting they are carriers but that this is not the cause of their presentation.

**Table 2:** Repeat expansion variant findings.

| Individual ID | Sex | Age at testing | Age at onset | FHx of CA | Gene | Disease | Inheritance | Variant motif | EH5 RE size | size | PCR RE size | Zygosity | Clinical presentation |
| --- | --- | --- | --- | --- | --- | --- | --- | --- | --- | --- | --- | --- | --- |
| P66 | F | 5th Decade | 5th Decade | - | ATXN8/ATXN8OS | SCA8 | AD | (CAG)<sub>n</sub>/(CTG)<sub>n</sub> | 100 | 124.2 | 134 | Het | CA and migraines |
| P110 | F | 9th Decade | 8th Decade | - | ATXN8/ATXN8OS | SCA8 | AD | (CAG)<sub>n</sub>/(CTG)<sub>n</sub> | 150 | 768.4 | Expanded^ | Het | CABV |
| P3 | M | 8th Decade | 7th Decade | - | FGF14 | SCA27B | AD | (GAA)<sub>n</sub> | 134 | 288.5 | 286 | Het | CABV, ANS dysfunction and cough |
| P4 | F | 8th Decade | 6th Decade | - | FGF14 | SCA27B | AD | (GAA)<sub>n</sub> | 105 | 327.1* | 346 | Het | CA and cough |
| P5 | M | 8th Decade | 6th Decade | - | FGF14 | SCA27B | AD | (GAA)<sub>n</sub> | 101 | 328.3* | 308 | Het | hyperreflexia |
| P13 | M | 7th Decade | 5th Decade | - | FGF14 | SCA27B | AD | (GAA)<sub>n</sub> | 126 | 445.5* | 447 | Het | CA |
| P14 | M | 8th Decade | 8th Decade | - | FGF14 | SCA27B | AD | (GAA)<sub>n</sub> | 150 | 260.9 | 310 | Het | CABV, hyperreflexia and parkinsonism |
| P22 | M | 7th Decade | ND | - | FGF14 | SCA27B | AD | (GAA)<sub>n</sub> | 114 | 108.3*~ | >400 | Het | CA and spasticity |
| P25 | F | 8th Decade | 5th Decade | + | FGF14 | SCA27B | AD | (GAA)<sub>n</sub> | 118 | 311.8 | 315 | Het | CABV and cough |
| P26 | F | 8th Decade | 7th Decade | - | FGF14 | SCA27B | AD | (GAA)<sub>n</sub> | 138 | 16.2*~ | 293 | Het | CA |
| P38 | M | 6th Decade | 5th Decade | - | FGF14 | SCA27B | AD | (GAA)<sub>n</sub> | 95 | 324.9* | 313 | Het | CA and spasticity |
| P40 | F | 7th Decade | 6th Decade | - | FGF14 | SCA27B | AD | (GAA)<sub>n</sub> | 188 | 126.8* | >400 | Het | hyperreflexia |
| P42 | F | 9th Decade | 6th Decade | - | FGF14 | SCA27B | AD | (GAA)<sub>n</sub> | 126 | 252.8 | 253 | Het | CABV |
| P45 | M | 8th Decade | ND | - | FGF14 | SCA27B | AD | (GAA)<sub>n</sub> | 145 | 329.6 | 349 | Het | CABV |
| P50 | M | 8th Decade | 7th Decade | - | FGF14 | SCA27B | AD | (GAA)<sub>n</sub> | 130 | 270.6* | 265 | Het | CABV |
| P52 | F | 9th Decade | 8th Decade | - | FGF14 | SCA27B | AD | (GAA)<sub>n</sub> | 117 | 277.3* | 281 | Het | CA |
| P63 | F | 8th Decade | ND | + | FGF14 | SCA27B | AD | (GAA)<sub>n</sub> | 119 | 325.4* | 327 | Het | CA and Hashimoto's thyroiditis |
| P64 | F | 8th Decade | 8th Decade | - | FGF14 | SCA27B | AD | (GAA)<sub>n</sub> | 182 | 255.7* | 287 | Het | CA |
| P67 | M | 9th Decade | 7th Decade | - | FGF14 | SCA27B | AD | (GAA)<sub>n</sub> | 186 | 265.2 | 267 | Het | CABV |
| P68 | M | 8th Decade | 8th Decade | - | FGF14 | SCA27B | AD | (GAA)<sub>n</sub> | 133 | 249.1* | 268 | Het | CA |
| P72 | M | 6th Decade | 6th Decade | + | FGF14 | SCA27B | AD | (GAA)<sub>n</sub> | 146 | 288.3 | 289 | Het | EA |
| P90 | M | 8th Decade | 8th Decade | + | FGF14 | SCA27B | AD | (GAA)<sub>n</sub> | 109 | 310.4 | 315 | Het | CA |
| P100 | F | 8th Decade | 7th Decade | + | FGF14 | SCA27B | AD | (GAA)<sub>n</sub> | 90 | 378 | 374 | Het | CABV |
| P101 | M | 8th Decade | 6th Decade | + | FGF14 | SCA27B | AD | (GAA)<sub>n</sub> | 151 | 316.4 | 317 | Het | EA |
| P106 | F | 7th Decade | 7th Decade | + | FGF14 | SCA27B | AD | (GAA)<sub>n</sub> | 148 | 316.4 | 310 | Het | CA |
| P107 | M | 8th Decade | ND | + | FGF14 | SCA27B | AD | (GAA)<sub>n</sub> | 101 | 325.3 | 322 | Het | CA |
| P95 | M | 4th Decade | 2nd Decade | - | FXN | FRDA | AR | (GAA)<sub>n</sub> | 127 | 295.6 | Expanded# | Hom | CA and UMN signs |
| P23 | M | 7th Decade | 6th Decade | + | NOP56 | SCA36 | AD | (GGCCTG)<sub>n</sub> | 317 | 5.1* | Expanded^ | Het | CA |
| P69 | M | 8th Decade | ND | - | RFC1 | CANVAS | AR | (AAGGG)<sub>n</sub> | 84 | No call | Expanded^ | Hom | CANVAS |
| P78 | M | 8th Decade | ND | - | RFC1 | CANVAS | AR | (AAGGG)<sub>n</sub> | 61 | 872.4~ | Expanded^ | Hom | CANVAS |
| P79 | F | 8th Decade | ND | - | RFC1 | CANVAS | AR | (AAGGG)<sub>n</sub> | 44 | 794~ | Expanded^ | Hom | CANVAS |
| P93 | M | 7th Decade | 7th Decade | - | RFC1 | CANVAS | AR | (AAGGG)<sub>n</sub> | 60 | 944.8~ | Expanded^ | Hom | CANVAS |
| P97 | M | 9th Decade | 6th Decade | - | RFC1 | CANVAS | AR | (AAGGG)<sub>n</sub> | 48 | No call | Expanded^ | Hom | CANVAS |

\* less than 7 supporting reads.
~ only one allele size called at this loci.

Overall, this analysis identified 46 individuals with potentially pathogenic RE causing their clinical presentation, including heterozygous RE in *ATXN8/ATXN8OS* (2x), *NOP56* (1x) and *FGF14* (37x), and biallelic RE in *FXN* (1x) and *RFC1* (5x).

### Molecular validation of RE identified by SR-GS

We and others have shown that SR-GS has high sensitivity and specificity for identification and sizing of pathogenic RE when the DNA repeat length is similar or smaller than the standard SR-GS read length of 150 base pairs (Dolzhenko et al. 2017; Tankard et al. 2018). In contrast, while SR-GS demonstrates high specificity/sensitivity for larger pathogenic RE, such as those causing SCA36 and myotonic dystrophy 2 (DM2), it significantly underestimates the size of the pathogenic allele (Day et al. 2003; Rafehi et al. 2020). We previously demonstrated that size estimates of the RE in *FGF14* that cause SCA27B are unreliable when the expansion is greater than ∼(GAA)_100_ (Rafehi et al. 2023), suggesting SR-GS may have poor specificity and/or sensitivity for this RE. Therefore, we performed orthogonal molecular testing of all individuals with potential pathogenic RE identified by SR-GS (*FXN* via clinical diagnostic test; *NOP56* and *RFC1* via repeat primed PCR; *ATXN8/ATXN8OS* and *FGF14* via repeat primed PCR and flanking PCR/capillary array sizing). This analysis confirmed the SR-GS results for the nine individuals with pathogenic RE in *ATXN8/ATXN8OS*, *FXN*, *NOP56*, and *RFC1* (Table 2). Of the 37 individuals with a potential pathogenic RE in *FGF14* only 24 were confirmed to be expanded above the current pathogenic threshold of (GAA)≥250 (Pellerin et al. 2023; Rafehi et al. 2023). We subsequently used long range PCR to size the *FGF14* locus in all individuals in the cohort, demonstrating significant divergence in the size of the larger allele estimated by EH5 compared to PCR (Figure 2C). These results demonstrate that SR-GS has a sensitivity of 100% and specificity of 85% to identify pathogenic RE in *FGF14* when utilizing an EH5 estimate of 90 repeats as the pathogenic threshold (Supplemental_Table_S2.xls). In addition, the PCR analysis identified nine individuals with an *FGF14* GAA allele greater than 179 but less than 250 repeats; these were classified as variants of uncertain significance (VUS) (Mohren et al. 2024) (Supplemental_Table_S3.xls). Overall, while SR-GS identified 46 individuals with a potentially pathogenic RE (42% of the cohort), the actual diagnostic rate achieved was 33/110 (30%), the discrepancy being the result of 13 false positive *FGF14* diagnoses (i.e. a GAA repeat less than 250 repeats).

### Identification of non-RE pathogenic variants with SR-GS

In addition to RE, a variety of other pathogenic variants can cause CA (Beaudin et al. 2019; Rudaks et al. 2024). Therefore, we analyzed the SR-GS data for SNV/indel, CNV and mitochondrial DNA (mtDNA) variants. No CNV or mtDNA variants were identified but an additional seven patients were solved by identification of likely pathogenic/pathogenic (LP/P) variants in *ANO10* (autosomal recessive spinocerebellar ataxia 10; SCAR10), *CACNA1G* (SCA42), *HEXA* (Tay Sachs disease), *PNPT1* (SCA25), *SPG7* (spastic paraplegia 7; SPG7), *STUB1* (SCA48), and *TTBK2* (SCA11) (Table 3). This analysis also identified an additional nine individuals with suspicious variants that do not meet ACMG guidelines for classification as LP/P (Richards et al. 2015) and were therefore scored as VUS (Supplemental_Table_S4.xls). The overall diagnostic yield achieved by SR-GS was 38% (33x RE, 7x non-RE), with a false positive/negative rate of 12% and 0% respectively for *FGF14* RE (SCA27B).

**Table 3.** Single nucleotide and small insertion or deletion variant findings.

| Individual ID | Sex | Age at testing | Age at onset | FHx of CA | Gene | Disease | Inheritance | Genomic variant (hg38) | HGVSc | HGVSp | Zygosity | Consequence | Classification | Clinical presentation |
| --- | --- | --- | --- | --- | --- | --- | --- | --- | --- | --- | --- | --- | --- | --- |
| P74 | F | 6th Decade | 4th Decade | - | *ANO10* | SCAR10 | AR | chr3:43574873T>C | c.1163-9A>G |  | Hom | splice acceptor | P | CA |
| P109 | M | 7th Decade | ND | + | *CACNA1G* | SCA42 | AD | chr17:50617560G>A | c.5144G>A | p.(Arg1715His) | Het | missense | P | CA |
| P18 | M | 4th Decade | 3rd Decade | - | *HEXA* | Tay Sachs disease | AR | chr15:72350518C>T | c.805G>A | p.(Gly269Ser) | Het | missense | P | CA and muscle weakness |
|  |  |  |  |  |  |  |  | chr15:72375740C>T | c.233G>A | p.(Trp78*) | Het | stop gained | P |  |
| P49 | F | 6th Decade | 2nd Decade | + | *PNPT1* | SCA25 | AD | chr2:55643216G>C | c.2014-3C>G |  | Het | splice acceptor | LP | CA |
| P96 | M | 8th Decade | 7th Decade | - | *SPG7* | SPG7 | AR | chr16:89546737C>T | c.1529C>T | p.(Ala510Val) | Hom | missense | P | CA, spasticity and osteoarthritis |
| P20 | M | 7th Decade | ND | - | *STUB1* | SCA48 | AD | chr16:681504CGAA>C | c.433_435del | p.(Lys145del) | Het | inframe deletion | LP | CA, akathisia and hyperreflexia |
| P59 | M | 8th Decade | 8th Decade | + | *TTBK2* | SCA11 | AD | chr15:42777132ATC>A | c.1306_1307del | p.(Asp436Tyrfs*14) | Het | frameshift | P | dementia |

M, male; F, female; ND, no data available; FHx, family history; CA, cerebellar ataxia; -, absent; +, present; SCAR10, autosomal recessive spinocerebellar ataxia type 10; SPG7, spastic paraplegia 7;
AD, autosomal dominant; AR, autosomal recessive; Het, heterozygous; Hom, homozygous; P, pathogenic; LP, likely pathogenic.
Transcripts: *ANO10* , NM\_018075.5; *CACNA1G* , NM\_018896.5; *HEXA* , NM\_000520.6; *PNPT1* , NM\_033109.5; *SPG7* , NM\_003119.4; *STUB1* , NM\_005861.4; *TTBK2* , NM\_173500.4.

### RE identification with long-read adaptive sequencing (LR-AS)

There is currently no way of coupling targeting with long read sequencing such that very deep sequencing of selected targets can be achieved at a reasonable cost. LR-AS is an attractive alternative which is cost effective and achievable. Therefore, LR-AS was performed targeting 334 genomic regions (111Mb/∼3.59% of the hg38 reference genome), which included 67 genes with a pathogenic RE and other non-RE genes associated with CA (Supplemental_Table_S5.xls). Each sample was analyzed using a single MinION flow cell. Summary statistics are presented in Figure 3 and detailed individual sample/locus results are described in Supplemental_Table_S5.xls. The on-target mean read length was 4,727 bp [ 95% CI 2,728-6,014] compared to 480 bp [95% CI 444, 493] for off-target sequence, confirming rapid and efficient rejection of non-targeted regions of the genome (Figure 3A). The on-target mean sequencing depth was 22.1 [95% CI 12.7, 31.0] compared to 3.0 [95% CI 1.7, 4.1] for off-target regions (Figure 3B), resulting in a mean 8.2-fold [95% CI 5.3-fold, 10.4-fold] target enrichment (Figure 3C). Consistent with the SR-GS analysis, we focused analysis on the 22 RE listed in Table 1. The distribution of read counts across all samples at these 22 loci (Figure 3D-E) ranged from a mean of 16.8 reads (*AR*) to 28.2 reads (*C9orf72*) across the 110 individuals in the cohort.

**Figure 3.**
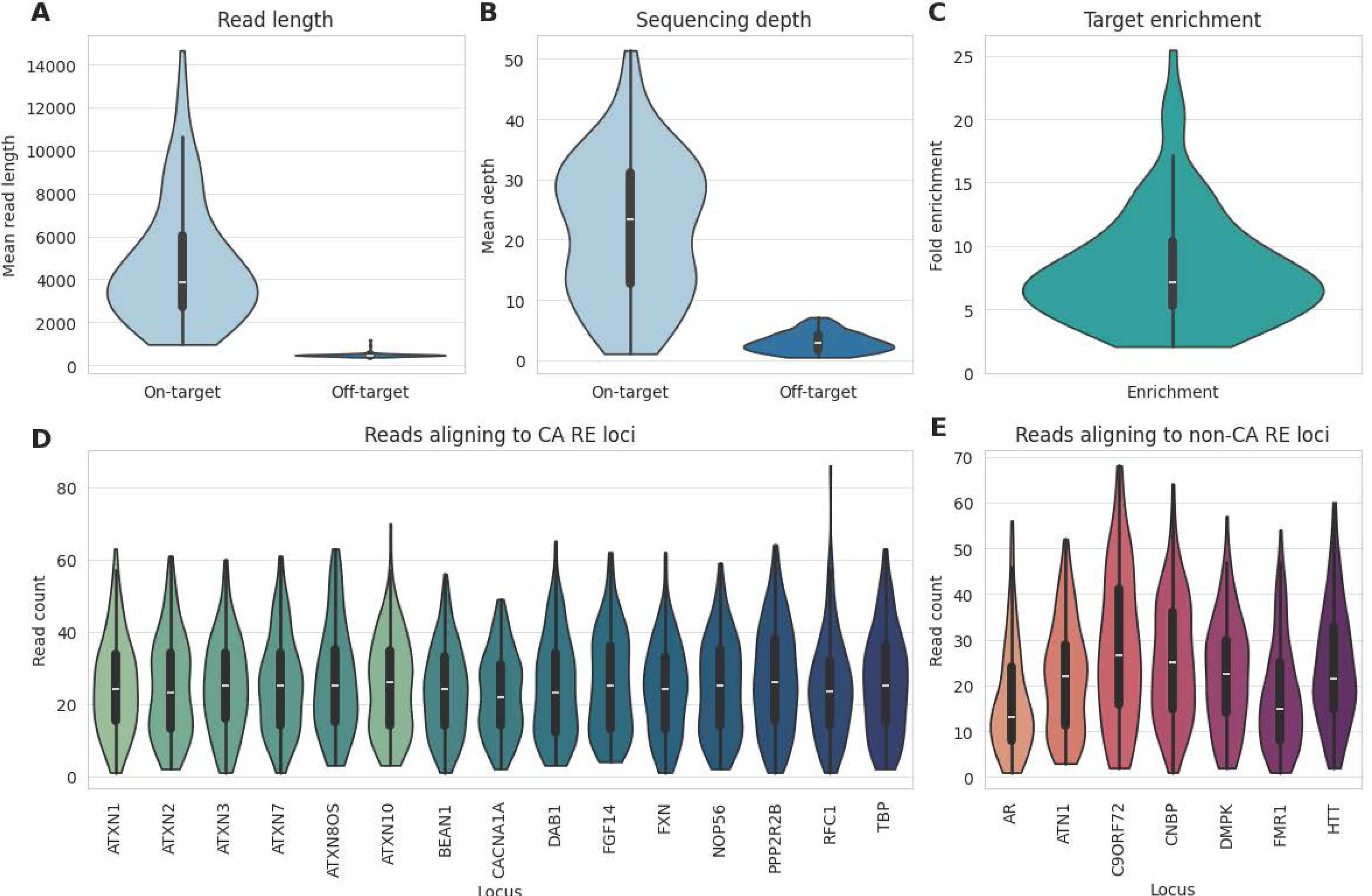
Performance metrics for adaptive sequencing in a targeted panel of repeat expansion loci. A. Mean read length in target regions significantly exceeds that of off-target regions. B. Sequencing depth in on-vs off-target regions. C. Genome-wide enrichment of on-target regions vs off-target regions in individual experiments. D. Number of reads aligning to each of the targeted cerebellar ataxia (CA) repeat loci. E. Number of reads aligning to each targeted clinically significant non-CA locus.

Given that clinical testing excluded individuals with SCA1/2/3/6 and 7 from the cohort, we first tested if LR-AS was effective in identifying the corresponding pathogenic RE by analyzing individuals with known pathogenic RE status. Affected individuals, not part of the cohort, with known pathogenic RE in *ATXN1*, *ATXN2*, *ATXN3*, *CACNA1A* and *ATXN7* were subject to LR-AS and genotypes were compared to allele sizes determined by diagnostic testing (flanking PCR and capillary array analysis). LR-AS identified two alleles, one pathogenic and one non-pathogenic, for *ATXN1, ATXN2, ATXN3* and *ATXN7*. The calculated allele sizes were concordant with those obtained by diagnostic testing (Supplemental_Table_S6.xls). However, Straglr only identified a single allele (14.4 repeats) for *CACNA1A*, compared to the diagnostic result of two alleles of 23 and 12 repeats. Visual inspection in IGV showed four spanning reads with 22-23 repeats, and 10 spanning reads with 12 repeats. It is not clear why Straglr was unable to call two alleles; it may be that the relatively low read depth and allelic bias (4 reads versus 10 reads) confounded the analysis. However, *ATXN7* was called successfully despite lower read depth and similar allelic bias (Supplemental_Table_S6.xls). Alternatively, the small relative difference in size between pathogenic and non-pathogenic alleles may have contributed to only a single allele being identified. While previous studies have suggested that LR-AS can effectively identify pathogenic RE in these five genes, they also highlighted the utility of manual inspection/visual confirmation of bioinformatic allele size estimations (Miyatake et al. 2022a; Stevanovski et al. 2022). This is a limitation that also impacts SR-GS as described above.

We subsequently extended the LR-AS analysis to the trial cohort, identifying individuals with potentially pathogenic RE by determining if their allele size estimates exceeded thresholds in a process similar to that used for SR-GS (Methods, Table 1). Overall, prior to any manual review Straglr identified RE that supported 30 potential diagnoses. For the dominant CA disorders, Straglr identified heterozygous pathogenic RE in *ATXN8OS* in two individuals (Figure 4). One RE was estimated to be 124 repeats, similar to the size estimated by orthogonal PCR analysis (134 repeats), while the second RE was estimated to be 768 repeats (Table 2), which is larger than can be determined by PCR. Heterozygous pathogenic RE in *FGF14* (≥250 GAA) were identified in 20 individuals and showed very high concordance with the PCR sizing results (R2=0.997, Figure 5A, Table 2). Expanded sequences were also detected for the benign reference motif (AAAAT) in *BEAN1* and *DAB1* (Figure 4A) but not the pathogenic motifs (TGGAA) or (ATTTC). Manual review of the aligned reads did not provide any evidence of pathogenic motifs embedded deep within the expanded sequence, which would have been missed with SR-GS (Supplemental_Fig_S1.pdf).

**Figure 4.**
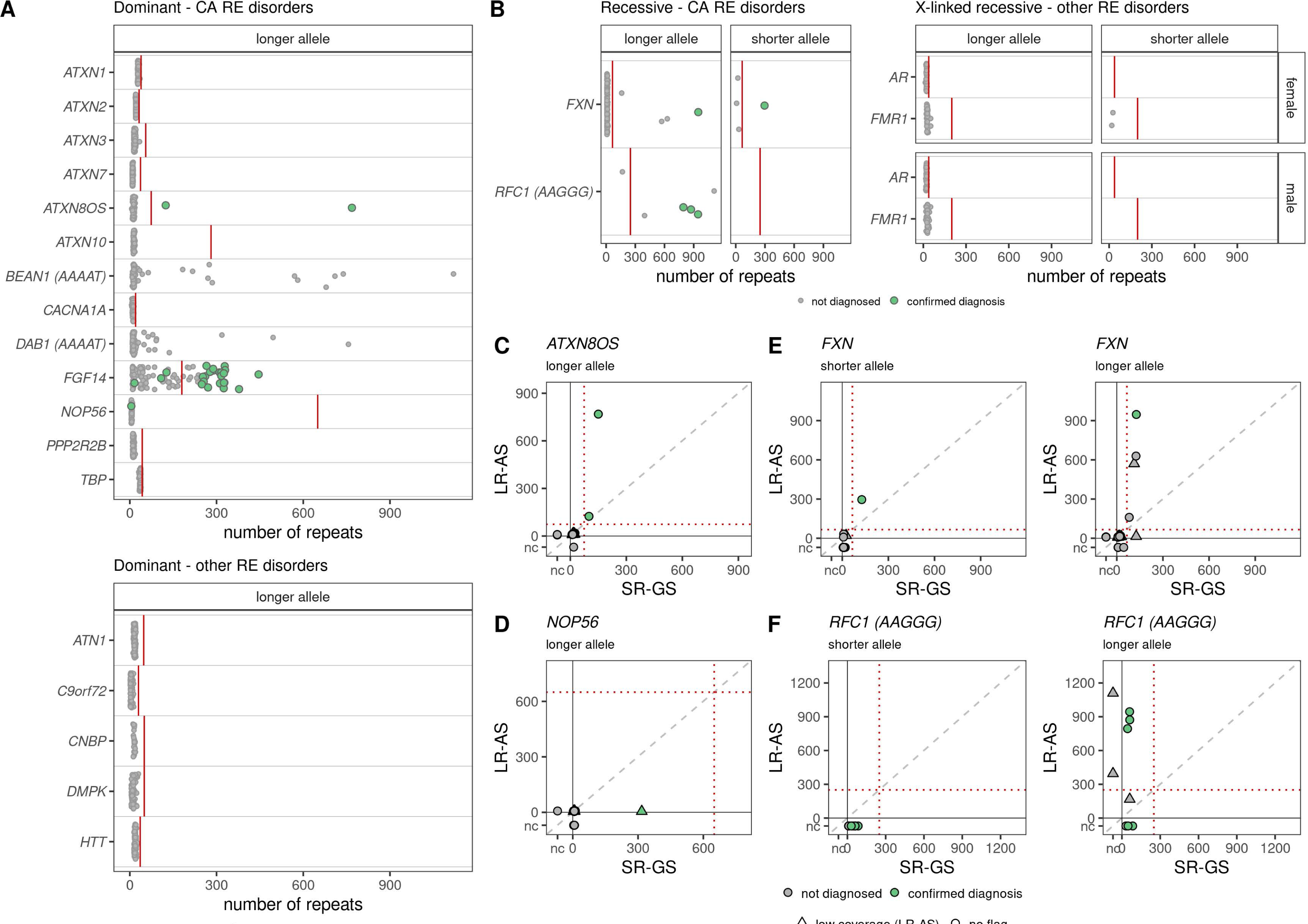
Targeted repeat expansion screening panel in LR-AS and comparison to SR-GW. STR genotypes were determined for short-listed loci with Straglr. Green points indicate individuals with a confirmed clinical diagnosis for the respective locus. A. Genotypes are shown for the longer allele in dominant RE disorders that cause ataxia (top) and those that cause other disorders (bottom). B. Genotypes for the shorter and longer alleles are shown for autosomal recessive RE ataxia disorders (left) and X-linked recessive disorders other than ataxia, split by sex (right). Comparison of allele genotyping with SR-GS and LR-GS is shown for loci with a confirmed diagnosis for the longer allele in dominant disorders. C. *ATXN8OS* (SCA8), D. *NOP56* (SCA36), and for both alleles in recessive disorders for E. *FXN* (FRDA) and F. *RFC1* (CANVAS, AAGGG motif only). LS-AS with low coverage (≤7 reads) are shown as triangles, those with no coverage issues flagged (>7 reads) are shown as circles.

**Figure 5.**
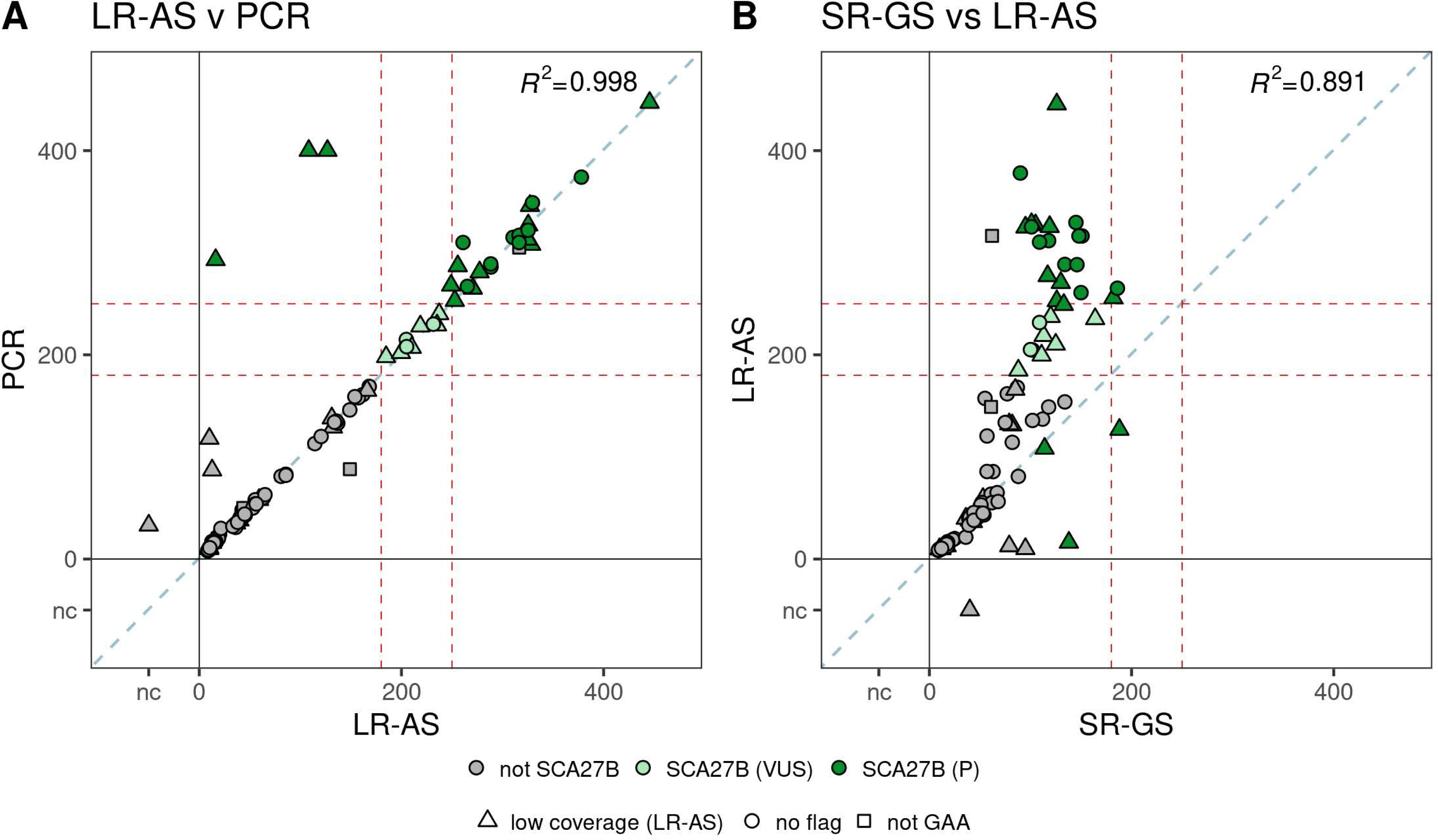
Comparison of *FGF14* STR sizing between LR-AS, SR-GS and PCR identifies strengths of LR-AS. Comparison of *FGF14* STR sizing is shown as concordance plots for A. LR-AS compared to PCR and B. LR-AS compared to SR-GS. The R-squared is a Pearson’s correlation. Dark green circles are individuals with a confirmed SCA27B diagnosis (P) and those in light green are SCA27B VUS. Triangles indicate low coverage (≤ 7 reads) on LR-AS, squares indicate non-GAA repeat motifs and circles samples with coverage >7 reads and GAA motifs. The pathogenic (≥250 repeats) and VUS (≥180 repeats) thresholds are shown as dashed red lines. A 1:1 correlation is shown as a dashed blue line. The R2 is a Pearson’s correlation. Individuals with no genotype call from either LR-AS, SR-GW or PCR are indicated separately to the numbered axis and are labelled nc (i.e. no genotype call).

Analysis of recessive loci also identified expanded RE in *FXN* and *RFC1*. Pathogenic *FXN* RE of 947 and 296 repeats were identified in one individual, confirming a molecular diagnosis of FRDA. Three other individuals were identified with a pathogenic GAA RE in *FXN*, with estimated sizes of 629, 570, and 160 repeats respectively (Figure 4B). However, the second allele for each individual was in the normal range (32, 21 and 8 repeats respectively), identifying these individuals as heterozygous carriers (Supplemental_Table_S1.xls). Pathogenic RE in *RFC1* were identified by Straglr in six individuals. Five of these were called ‘homozygous’ RE for the pathogenic AAGGG, i.e. only a single allele size was reported. The estimated allele sizes of 872, 794, 945, 395 and 1109 repeats exceed the current pathogenic threshold of ≥250 repeats (Dominik et al. 2023). Three of these were confirmed as biallelic expansions by visual review of aligned reads (Table 2), while the other two (395 and 1109 repeats) were shown to be heterozygous for a single pathogenic AAGGG RE. In both cases Straglr did not identify (call) the second non-pathogenic allele. On visual inspection in IGV, the samples appeared to be heterozygous in motif as well as size, with both samples having one pathogenic AAGGG allele, and a second AAAAG allele. Subsequent flanking PCR and gel electrophoresis confirmed the presence of a non-pathogenic AAAAG allele in both P10 and P5 (Supplemental_Fig_S2A-B.pdf). In addition, one individual was called ‘homozygous’ RE for the pathogenic AAAGG, with an estimated allele size of 640 repeats, exceeding the current pathogenic threshold of >500 repeats for this motif (Dominik et al. 2023). However, manual review determined that the motif is actually AAGGG. Additionally, Straglr did not call the second non-pathogenic 130 repeat AAAGG allele (Supplemental_Fig_S2C.pdf), therefore this individual is also a heterozygous carrier.

### Comparison of testing methods

Independent analysis of the cohort by LR-AS, SR-GS and for some loci orthogonal PCR methodologies enabled us to compare and contrast the results across the different platforms. Allele sizing was broadly concordant between LR-AS and SR-GS data up to a RE size of approximately 450 bp (Figure 4, Supplemental_Fig_S3). This observation is consistent with previously published work performing PCR validation of long-read RE sizing methods (Ibanez et al. 2022). Expansions in excess of 450 bp were consistently sized by LR-AS as being larger than SR-GS due to the longer read length afforded by the Oxford Nanopore Technologies (ONT) platform. There was a very good correlation between LR-AS and orthogonal sizing methodologies (Figure 4, Table 2), indeed LR-AS with read lengths >5kb actually exceeds the sizing capabilities of PCR/capillary array technology (limited to <2kb).

Straglr did not size the single sample with a pathogenic *NOP56* expansion (P23) due to a lack of any reads completely spanning the expanded allele. However, manual review clearly identified reads demonstrating the presence of a pathogenic heterozygous expansion at this locus (Supplemental_Fig_S4A.pdf). Similarly, two samples with pathogenic biallelic *RFC1* expansions (P69, P97 confirmed by SR-GS and PCR testing) were not identified by Straglr due to low read depth; in each case only a single read spanned the entire RE. However, manual review confirmed the majority of reads at this locus encoded the pathogenic AAGGG motif, suggesting the individuals had biallelic pathogenic *RFC1* RE (Supplemental_Fig_S4B-C.pdf).

Overall, manual review of LR-AS was an important step in correct interpretation and diagnoses of very large RE. Our results also demonstrate the need for orthogonal methodologies for accurate interpretation of *RFC1* alleles, which can present with significant size and motif heterogeneity. LR-AS identified 29 pathogenic RE and achieved a genetic diagnosis for 26% of the cohort. This included heterozygous RE in *ATXN8/ATXN8OS* (2x), *NOP56* (1x) and *FGF14* (20x), and biallelic RE in *FXN* (1x) and *RFC1* (5x). This is four less than the 33 pathogenic RE identified using SR-GS, all of which were individuals with SCA27B. In one of these four cases, LR-AS determined an allele size of 249 repeats, compared to 268 repeats by PCR. This falls just under the current pathogenic threshold and therefore was classified as a VUS. For the other three, the read depth was low (<7) and only a single allele (less than 250 repeats) was called by Straglr (Table 2).

## DISCUSSION

There are over 100 clinically recognized disorders encompassed by CA making genetic diagnosis challenging. CA can present with non-specific and overlapping clinical features, providing minimal guidance for prioritizing genetic candidates and optimal diagnostic methodologies. RE are the predominant genetic cause of CA but until recently they were difficult to diagnose with existing molecular tools. Therefore, CA testing has been fragmented, inefficient and often restricted to single or a small number of RE loci. Patients often endure a long diagnostic odyssey with sequential testing, frequently to no avail (Nemeth et al. 2013; Daker-White et al. 2015; Rexach et al. 2019; Ngo et al. 2020). This study compares two promising technologies for comprehensive genetic diagnosis of CA, focusing on identifying pathogenic RE.

The cohort characteristics are broadly consistent with expectations for adult-onset cerebellar disorders, with similar numbers of males and females. Symptom onset (56±14 years) considerably preceded mean age at testing (68±13 years), reflecting a slowly progressive disease course. Diagnoses included SCA8 (2x), SCA27B (24x), SCA36 (1x), CANVAS (5x) and FRDA (1x). Our data confirm RE as the most common cause of CA in Australia, even in a cohort depleted of five commonly tested RE (SCA1, 2, 3, 6 and 7). However, biases in cohort collection may have influenced disorder frequency compared to true Australian population prevalence. Individuals were predominantly recruited from a single centre and therefore may not be broadly representative. In addition, cohort recruitment was mediated by the clinician referring patients for diagnostic testing, introducing potential bias depending on clinician expertise and enthusiasm for research.

SR-GS achieved 40 diagnoses (40/110, 36% yield) including 33 pathogenic RE and seven other non-RE causes. One advantage of SR-GS is its maturity and well established pipelines for different types of genetic variation. It is most efficient in identifying small variants (SNV and indel) with LP/P variants accounting for 6% of the cohort. In addition, nine clinically suspicious variants were identified as likely candidates (VUS), requiring future clinical followup and functional studies to confirm pathogenicity. Overall clinically relevant small variants were found in 15% of cases, consistent with literature on their contribution to CA (Rudaks et al. 2024). Notably, all of these variants affected protein coding or nearby flanking sequences and therefore are detectable by standard exome sequencing.

Bioinformatic tools capable of identifying pathogenic RE in SR-GS data are a relatively recent development, and have yet to be widely embraced in diagnostic pipelines (Bahlo et al. 2018; Leitao et al. 2024). This study demonstrates SR-GS’s utility in identifying pathogenic RE causing CA. Analysis of 17 RE loci associated with CA yielded 33 genetic diagnoses, including both dominant and recessive disorders. Tools like EH5 estimate STR sizes and can utilize reported pathogenic thresholds for RE, such as *ATXN8OS*, which are smaller than the read length. However, larger pathogenic RE require empirical establishment of thresholds that maximize the specificity and sensitivity of RE identification. This is straightforward for disorders such as CANVAS, where a non-reference motif causes disease. In our experience detection of 10 or more non-reference pathogenic motifs provides strong suspicion of a pathogenic RE. Non-reference motif conditions like SCA31 and SCA37 present challenges as pathogenic motifs may be embedded within an expanded non-pathogenic motifs (Rosenbohm et al. 2022). Thousands of alleles in gnomAD have non-pathogenic reference motif (AAAAT) expansions >30 repeats for SCA31 and SCA37 (out of 18,511 individuals). SR-GS methods are mostly unable to detect cases where a pathogenic RE is embedded deep within non-pathogenic motifs due to short DNA fragment libraries and reliance on sequencing reads aligning to non-repetitive regions adjacent to a putative RE

Our findings show that SCA27B, caused by pathogenic expansion of the reference GAA motif, is potentially the most common genetic cause of adult-onset ataxia in Australia. Although no diagnostic test is currently available in Australia for this condition, our results demonstrate SR-GS’s effectiveness as a screening tool. Despite considerable population variability in non-pathogenic GAA allele size, applying an empirical threshold of >90 repeats using EH5 provides optimal sensitivity (100%) with an acceptable false positive rate (16%). These outcomes are consistent with an independent retrospective cohort study, which showed good predictive value (70%) and excellent sensitivity (100%) using STRling for allele size estimates (Mohren et al. 2024). Both studies highlight the need for follow-up targeted orthogonal analysis to accurately determine RE size in cases identified by SR-GS, especially given SCA27B’s recent identification and uncertainty regarding pathogenic thresholds. While the original studies suggest a pathogenic threshold ≥250 repeats (Pellerin et al. 2023; Rafehi et al. 2023), subsequent studies suggest higher (>300 (Mereaux et al. 2024)) or lower (>180 (Mohren et al. 2024)) thresholds. We identified nine individuals (8% of the cohort) with *FGF14* GAA alleles in the 180-249 range and report them as VUS. No alternative pathogenic variants were found in these cases, providing additional support that this range may be pathogenic.

The second most common cause of CA identified in our cohort was biallelic RE in *RFC1* (5/110, 5% yield). Our results demonstrate SR-GS’s ability to prioritize cases with potential pathogenic RE at this locus, but extensive orthogonal testing, expert variant interpretation and clinical input were essential for accurate *RFC1* disorder diagnosis. Awareness of diverse pathogenic motifs and thresholds for this disorder is growing, presenting challenges for effective molecular diagnosis utilizing SR-GS (Miyatake et al. 2022b; Scriba et al. 2023). While additional bioinformatic tools can address some issues (Sullivan et al. 2024) SR-GS will likely function best as a pre-screening method for *RFC1* diagnostics. A long-read diagnostic tool capable of haplotype-resolved read alignments and full motif composition interrogation will be needed for effective genetic analysis and interpretation of this locus.

LR-AS is a nanopore-based long read sequencing application that enables *in silico* target selection in real time and has tremendous diagnostic potential. Advantages include prioritizing of likely disease-causing loci/variants, reducing curation time and improving resource utilization by enabling higher sequence multiplexing. In RE disorders, including CA, sequencing depth is critical as knowledge of RE size and composition affects disease prognosis and management (Hannan 2018; Consortium 2019). This study achieved a mean ∼eight-fold target enrichment and on-target mean sequencing depth of 22 (95% CI 12.7, 31.0, Figure 3). This was insufficient for local assembly of diploid alleles and motif composition interrogation for a majority (75%) of the 2420 RE (22 x 110) targeted. Our results contrast with reports of successful haplotype-resolved allele interrogation with similar metrics (∼5x enrichment, ∼8-40x target coverage, (Miller et al. 2021; Stevanovski et al. 2022)). Reliable and reproducible data are essential for accreditation of a diagnostic test, and it is possible these divergent results are due to the choice of bioinformatic tools implemented by the different studies. A current limitation of LR-AS and indeed the Oxford Nanopore Technologies (ONT) platform more broadly, is the lack of widely-accepted best-practice pipelines for bioinformatic analysis of data. The Epi2Me Labs wf-human-variation pipeline produced by ONT’s Customer Workflows group is the closest analogue to the GATK best practices broadly used in processing short-read sequencing data. We designed our analysis after the Epi2Me pipeline, using the most recent, highest-accuracy basecalling model available, and selecting minimap2 as recommended by recent benchmarks of alignment tools (LoTempio et al. 2023; Helal et al. 2024). Similarly, we selected Straglr (Chiu et al. 2021) for its compatibility with minimap2-aligned data, as it is the currently recommended STR caller in Epi2me’s human variation pipeline.

Despite read depth limitations, LR-AS performed exceptionally well in this trial, achieving 29 diagnoses (29/110, 26% yield). Greater manual review and interpretation of LR-AS data were required to support a diagnosis compared to SR-GS. For example, Straglr analysis did not identify potential RE in *NOP56*, despite 4/8 reads indicating a likely presence pathogenic RE (Supplemental_Fig_S4.pdf). Similarly, while Straglr identified six individuals with potential biallelic pathogenic RE in *RFC1*, extensive manual curation and orthogonal testing showed that only three had true biallelic RE. Moreover, Straglr was unable to generate an allele size estimate for two individuals that SR-GS and orthogonal analysis identified as biallelic. Pathogenic RE were identifiable in aligned reads, but low read depth and few spanning reads compromised allele identification and sizing.

RE identification by LR-AS was concordant with SR-GS, except for four individuals with SCA27B (Table 2). These diagnoses were missed due to low read depth at the *FGF14* loci, and do not constitute a systemic issue with interrogation of this loci. Indeed, LR-AS appears to have significant advantages over SR-GS in SCA27B diagnosis. No false positive results were observed and long-read sequencing can easily span the largest *FGF14* alleles reported to date (>900 repeats, (Mohren et al. 2024)) using current diagnostic workflows for gDNA extraction. High correlation in repeat length estimates with LR-PCR (Figure 5, R2=0.998) suggests orthogonal validation won’t be required when suitable read depth is achieved consistently.

Read depth is a major technical impediment to diagnostic implementation of LR-AS. While there is a lack of consensus in the field, a read depth of ∼33x supports phased *de novo* genome assembly (Porubsky et al. 2021). For diagnostic purposes 33x is likely the lower bound; ideally all would be spanning reads to support accreditation benchmarks for consistent and accurate characterization of RE (and small variants). Reliably achieving these targets may be challenging given the competing need to minimize the target panel for maximum enrichment versus increasing panel size to reduce nanopore destruction due to polarity reversal associated with off-target rejection. Additional enrichment strategies, including CRISPR/Cas9, have been proposed with LR-AS, or indeed as an alternative enrichment protocol (Mizuguchi et al. 2021; Lopatriello et al. 2023; Leitao et al. 2024). However, in the context of diagnostic application these approaches are bespoke, require considerable optimization and are limited in the number of targets that can be successfully multiplexed.

A major point of difference between SR-GS and LR-AS is the diversity of mutation types that can be interrogated. We used existing diagnostic grade pipelines for identification of small variants, CNV and alterations in mtDNA in the SR-GS data, showing pathogenic small variants as the second most common CA mutation class. We did not formally interrogate the AS-LR data for small variants given the read depth achieved and ONT sequence data error rates (Ni et al. 2023). However, subsequent visual inspection of aligned reads spanning pathogenic small variants identified by SR-GS revealed seven of eight variants were identifiable in the ONT reads, suggesting that LR-AS data analysis for RE and small variants could improve diagnostic yield (Supplemental_Fig._S5.png).

In conclusion, diagnosing CA remains challenging due to its complexity and variability. This study compared short-read genome sequencing (SR-GS) and long-read adaptive sequencing (LR-AS) for their efficacy in identifying pathogenic repeat expansions (RE), the predominant genetic cause of CA. SR-GS demonstrated a higher diagnostic yield (36%) and benefits from its mature, established clinical use, capable of identifying various genetic variants. It also supports future reanalysis as new RE and CA-associated genes are identified. However, its reliance on orthogonal validation for certain RE and limitations in mapping large, complex RE limit its use primarily as a screening tool. Conversely, LR-AS showed promise with a 26% diagnostic yield and potential for more precise haplotype resolution and targeted analysis of clinically relevant regions. Despite requiring extensive manual data interpretation and facing challenges with read depth and error rates, LR-AS has the potential to develop into a standalone frontline diagnostic test for RE. Both technologies represent significant advancements in CA diagnostics that require continued investment in technological development and computational resources. Cost will also significantly impact clinical implementation. SR-GS capacity is established in many diagnostic facilities but reagent costs per test are unlikely to decrease significantly. SR-AS will require investment to establish equipment and computational infrastructure but reagent costs per test could decrease due to sample multiplexing and targeted array analysis. Ultimately, a substantial proportion of the cost for both technologies lies in variant curation and interpretation. The future integration of enhanced variant interpretation algorithms, including artificial general intelligence capability, into diagnostic pipelines will benefit both platforms.

## Data Availability

All data produced in the present study are available upon reasonable request to the authors

## METHODS

### Cohort recruitment and clinical phenotype

The Royal Children’s Hospital Human Research Ethics Committee (HREC #28097) and the Walter and Eliza Hall Institute of Medical Research (HREC 18/06) approved the study. Multiple health practitioners in Australia referred individuals with a clinical diagnosis of CA for standard diagnostic testing of five RE loci (SCA1/2/3/6/7), mostly to a single center [VCGS]. Informed consent for research genetic testing was obtained from 110 participants undergoing clinical testing and phenotype details were derived from the pathology test request form or the referring clinician. Blood-derived genomic DNA was isolated by VCGS using standard methods and integrity confirmed by TapeStation (Agilent Technologies) analysis.

### Short-read genome sequencing and analysis

Genome sequencing was performed by the VCGS in Melbourne, Australia with the TruSeq PCR-free DNA HT Library Preparation Kit and sequenced on the Illumina NovaSeq 6000 platform at targeted mean coverage of 30x. For detection of pathogenic non-RE variants, data were bioinformatically processed by VCGS using commercially available and in-house analysis pipelines. Alignment to the reference genome (GRCh38) and calling of nuclear/germline DNA variants was performed using the Dragen v.3.3.7 (Illumina) workflow. Alignment to the revised Cambridge Reference Sequence (rCRS) mitochondrial genome (NC_012920.1) and calling of mtDNA variants was performed using an in-house analysis pipeline based on the Broad Institute best practice workflow. We used a research instance of seqr (Pais et al. 2022) implemented by Murdoch Children’s Research Institute, Melbourne, Australia, to annotate and filter SNV and indel variants. Variants were prioritized for curation using the PanelApp Australia Ataxia_Superpanel gene list (Version 3.13). Candidate variants were reviewed at a multidisciplinary team (MDT) meeting comprised of clinicians, genomic laboratory staff and bioinformaticians and classified based on ACMG guidelines (Richards et al. 2015). CNV were screened for and interpreted using an internal CNV detection tool, CXGo (Sadedin et al. 2018). For mtDNA variant interpretation, a custom VCGS analysis pipeline was used to visualize large deletions and annotate the VCF file with variant information before manual filtering. An mtDNA variant was regarded as homoplasmic or apparently homoplasmic when it was present in at least 97%, respectively, of sequence reads aligned to the genomic position. For RE detection, alignment and variant calling was performed according to the GATK best practice pipeline and analysis of 67 RE loci was performed utilizing exSTRa (v.1.1.0) (Tankard et al. 2018) and ExpansionHunter (v.5.0.0) (Dolzhenko et al. 2019) as previously described (Rafehi et al. 2023).

### Targeted long-read sequencing and analysis

Long-read sequencing was performed on Oxford Nanopore Technologies (ONT) MinION Mk1B sequencer. Nanopore sequencing libraries were prepared from ∼3 μg of gDNA, using SQK-LSK110 Ligation Sequencing Kit (Oxford Nanopore Technologies) and the NEBNext Companion Module (New England BioLabs) according to the manufacturer’s specifications. Samples were loaded onto FLO-MIN106D flow cells (Oxford Nanopore Technologies) and run for 16-20 hrs before performing a wash and reload using the Flow Cell Wash Kit (EXP-WSH004, Oxford Nanopore Technologies) and run for an additional 24-36 hrs. Targeted sequencing was performed using the Readfish software package (Payne et al. 2021). A panel of 334 regions of genome sequence were targeted, including 67 genes with a pathogenic RE associated with neurological/neuromuscular conditions, genes identified in the Ataxia_Superpanel (PanelApp Australia Version 3.13) and other candidate genes potentially associated with cerebellar ataxia. A full list of targets is provided in Supplemental_Table_S4.xls.

Libraries were processed using the wf-human-variation pipeline (Oxford Nanopore Technologies, v1.8.3), with super-accuracy basecalling model dna_r9.4.1_e8_sup from Dorado v0.3.1 on nVidia A100 and A30 GPUs. Minimum depth for the pipeline was set at 2x coverage. Reads were aligned to GRCh38 (’GRCh38_full_analysis_set_plus_decoy_hla’) using minimap2 (v2.24-r1122). Short tandem repeats were typed against a custom locus catalog containing 154 locus-motif pairs at 67 unique loci using Straglr (v1.4.1). Loci were manually reviewed with IGV (v.2.16.2).

### Molecular genetic studies

*FGF14* LR-PCR and RP-PCR were performed as previously described (Rafehi et al. 2023), but using LongAmp HotStart Taq (New England Biolabs). *RFC1* flanking PCR was performed as previously described (Rafehi et al. 2019). *RFC1* RP-PCR and LR-PCR were performed using Q5® High-Fidelity PCR Master Mix (New England Biolabs) and previously published primers (Cortese et al. 2019) *ATXN8/ATX8OS* sizing PCR and RP-PCR were performed as previously described (Zhou et al. 2019). *NOP56* RP-PCR was performed as previously described (Garcia-Murias et al. 2012). For all PCRs, fragment analysis of FAM labeled PCR products was performed on capillary array (ABI3730xl DNA Analyzer, Applied Biosystems) with a LIZ1200 size standard, and visualized using PeakScanner 2 (Applied Biosystems).

### Statistical analyses

Pearson’s correlation was performed in R (version 4.0.5) was used to determine concordance for *FGF14* size estimates between SR-GS, LR-AS and PCR. RE consisting of non-AAG motifs and low coverage LR-AS calls were excluded from the calculation. Sensitivity and specificity for FGF14 for SR-GS and LR-AS were calculated only for individuals with pure GAA motifs. For LR-GS, samples with read depth ≤7 were excluded from the calculations.

## DATA ACCESS

The supplementary data file consists of five figures and six tables. Sequencing data generated in this study will be submitted to the European Genome-phenome Archive (EGA) (https://ega-archive.org/).

## ACKNOWLEDGMENTS

This work was supported by the Australian Government National Health and Medical Research Council grants (GNT2001513 and MRFF2007677) to PJL, MBD, HR, MB and DS. HR was supported by a NHMRC Emerging Leadership 1 grant (1194364) and MB was supported by an NHMRC Investigator grant (1195236). Additional funding was provided by the Independent Research Institute Infrastructure Support Scheme, the Victorian State Government Operational Infrastructure Program and the Murdoch Children’s Research Institute. KRK reports financial support was provided by the Ainsworth 4 Foundation unrelated to the current study.

## REFERENCES

Ashley EA. 2016. Towards precision medicine. Nat Rev Genet 17: 507–522.

Bahlo M, Bennett MF, Degorski P, Tankard RM, Delatycki MB, Lockhart PJ. 2018. Recent advances in the detection of repeat expansions with short-read next-generation sequencing. F1000Res 7.

Beaudin M, Matilla-Duenas A, Soong BW, Pedroso JL, Barsottini OG, Mitoma H, Tsuji S, Schmahmann JD, Manto M, Rouleau GA et al. 2019. The Classification of Autosomal Recessive Cerebellar Ataxias: a Consensus Statement from the Society for Research on the Cerebellum and Ataxias Task Force. Cerebellum 18: 1098–1125.

Chiu R, Rajan-Babu IS, Friedman JM, Birol I. 2021. Straglr: discovering and genotyping tandem repeat expansions using whole genome long-read sequences. Genome biology 22: 224.

Consortium GMoHsDG-H. 2019. CAG Repeat Not Polyglutamine Length Determines Timing of Huntington’s Disease Onset. Cell 178: 887–900 e814.

Cortese A, Beecroft SJ, Facchini S, Curro R, Cabrera-Serrano M, Stevanovski I, Chintalaphani S, Gamaarachchi H, Weisburd B, Folland C et al. 2023. A CCG expansion in *ABCD3* causes oculopharyngodistal myopathy in individuals of European ancestry. medRxiv doi:10.1101/2023.10.09.23296582: 2023.2010.2009.23296582.

Cortese A, Simone R, Sullivan R, Vandrovcova J, Tariq H, Yan YW, Humphrey J, Jaunmuktane Z, Sivakumar P, Polke J et al. 2019. Biallelic expansion of an intronic repeat in RFC1 is a common cause of late-onset ataxia. Nature genetics 51: 649–658.

Daker-White G, Kingston H, Payne K, Greenfield J, Ealing J, Sanders C. 2015. ’You don’t get told anything, they don’t do anything and nothing changes’. Medicine as a resource and constraint in progressive ataxia. Health Expect 18: 177–187.

Day JW, Ricker K, Jacobsen JF, Rasmussen LJ, Dick KA, Kress W, Schneider C, Koch MC, Beilman GJ, Harrison AR et al. 2003. Myotonic dystrophy type 2: molecular, diagnostic and clinical spectrum. Neurology 60: 657–664.

de Silva R, Greenfield J, Cook A, Bonney H, Vallortigara J, Hunt B, Giunti P. 2019. Guidelines on the diagnosis and management of the progressive ataxias. Orphanet journal of rare diseases 14: 51.

Depienne C, Mandel JL. 2021. 30 years of repeat expansion disorders: What have we learned and what are the remaining challenges? American journal of human genetics 108: 764–785.

Dolzhenko E, Bennett MF, Richmond PA, Trost B, Chen S, van Vugt J, Nguyen C, Narzisi G, Gainullin VG, Gross AM et al. 2020. ExpansionHunter Denovo: a computational method for locating known and novel repeat expansions in short-read sequencing data. Genome biology 21: 102.

Dolzhenko E, Deshpande V, Schlesinger F, Krusche P, Petrovski R, Chen S, Emig-Agius D, Gross A, Narzisi G, Bowman B et al. 2019. ExpansionHunter: a sequence-graph-based tool to analyze variation in short tandem repeat regions. Bioinformatics 35: 4754–4756.

Dolzhenko E, van Vugt J, Shaw RJ, Bekritsky MA, van Blitterswijk M, Narzisi G, Ajay SS, Rajan V, Lajoie BR, Johnson NH et al. 2017. Detection of long repeat expansions from PCR-free whole-genome sequence data. Genome research 27: 1895–1903.

Dominik N, Magri S, Curro R, Abati E, Facchini S, Corbetta M, Macpherson H, Di Bella D, Sarto E, Stevanovski I et al. 2023. Normal and pathogenic variation of RFC1 repeat expansions: implications for clinical diagnosis. Brain 146: 5060–5069.

Fearnley LG, J. R, Bahlo M. 2024. EH5_kmerfilter v1.0.0. doi:doi.org/10.5281/zenodo.11514479.

Gall-Duncan T, Sato N, Yuen RKC, Pearson CE. 2022. Advancing genomic technologies and clinical awareness accelerates discovery of disease-associated tandem repeat sequences. Genome research 32: 1–27.

Garcia-Foncillas J, Argente J, Bujanda L, Cardona V, Casanova B, Fernandez-Montes A, Horcajadas JA, Iniguez A, Ortiz A, Pablos JL et al. 2021. Milestones of Precision Medicine: An Innovative, Multidisciplinary Overview. Mol Diagn Ther 25: 563–576.

Garcia-Murias M, Quintans B, Arias M, Seixas AI, Cacheiro P, Tarrio R, Pardo J, Millan MJ, Arias-Rivas S, Blanco-Arias P et al. 2012. ’Costa da Morte’ ataxia is spinocerebellar ataxia 36: clinical and genetic characterization. Brain 135: 1423–1435.

Hadjivassiliou M, Martindale J, Shanmugarajah P, Grunewald RA, Sarrigiannis PG, Beauchamp N, Garrard K, Warburton R, Sanders DS, Friend D et al. 2017. Causes of progressive cerebellar ataxia: prospective evaluation of 1500 patients. J Neurol Neurosurg Psychiatry 88: 301–309.

Hannan AJ. 2018. Tandem repeats mediating genetic plasticity in health and disease. Nat Rev Genet 19: 286–298.

Helal AA, Saad BT, Saad MT, Mosaad GS, Aboshanab KM. 2024. Benchmarking long-read aligners and SV callers for structural variation detection in Oxford nanopore sequencing data. Sci Rep 14: 6160.

Ibanez K, Polke J, Hagelstrom RT, Dolzhenko E, Pasko D, Thomas ERA, Daugherty LC, Kasperaviciute D, Smith KR, Group WGSfND, et al. 2022. Whole genome sequencing for the diagnosis of neurological repeat expansion disorders in the UK: a retrospective diagnostic accuracy and prospective clinical validation study. Lancet Neurol 21: 234–245.

Jayadev S, Bird TD. 2013. Hereditary ataxias: overview. Genet Med 15: 673–683.

Kang C, Liang C, Ahmad KE, Gu Y, Siow SF, Colebatch JG, Whyte S, Ng K, Cremer PD, Corbett AJ et al. 2019. High Degree of Genetic Heterogeneity for Hereditary Cerebellar Ataxias in Australia. Cerebellum 18: 137–146.

La Spada AR, Wilson EM, Lubahn DB, Harding AE, Fischbeck KH. 1991. Androgen receptor gene mutations in X-linked spinal and bulbar muscular atrophy. Nature 352: 77–79.

Lappalainen T, Scott AJ, Brandt M, Hall IM. 2019. Genomic Analysis in the Age of Human Genome Sequencing. Cell 177: 70–84.

Lee A. 2023. Omaveloxolone: First Approval. Drugs 83: 725–729.

Leitao E, Schroder C, Depienne C. 2024. Identification and characterization of repeat expansions in neurological disorders: Methodologies, tools, and strategies. Rev Neurol (Paris) 180: 383–392.

Lopatriello G, Maestri S, Alfano M, Papa R, Di Vittori V, De Antoni L, Bellucci E, Pieri A, Bitocchi E, Delledonne M et al. 2023. CRISPR/Cas9-Mediated Enrichment Coupled to Nanopore Sequencing Provides a Valuable Tool for the Precise Reconstruction of Large Genomic Target Regions. Int J Mol Sci 24.

LoTempio J, Delot E, Vilain E. 2023. Benchmarking long-read genome sequence alignment tools for human genomics applications. PeerJ 11: e16515.

McCarthy JJ, McLeod HL, Ginsburg GS. 2013. Genomic medicine: a decade of successes, challenges, and opportunities. Science translational medicine 5: 189sr184.

Mereaux JL, Davoine CS, Pellerin D, Coarelli G, Coutelier M, Ewenczyk C, Monin ML, Anheim M, Le Ber I, Thobois S et al. 2024. Clinical and genetic keys to cerebellar ataxia due to FGF14 GAA expansions. EBioMedicine 99: 104931.

Miller DE, Sulovari A, Wang T, Loucks H, Hoekzema K, Munson KM, Lewis AP, Fuerte EPA, Paschal CR, Walsh T et al. 2021. Targeted long-read sequencing identifies missing disease-causing variation. American journal of human genetics 108: 1436–1449.

Miyatake S, Koshimizu E, Fujita A, Doi H, Okubo M, Wada T, Hamanaka K, Ueda N, Kishida H, Minase G et al. 2022a. Rapid and comprehensive diagnostic method for repeat expansion diseases using nanopore sequencing. NPJ Genom Med 7: 62.

Miyatake S, Yoshida K, Koshimizu E, Doi H, Yamada M, Miyaji Y, Ueda N, Tsuyuzaki J, Kodaira M, Onoue H et al. 2022b. Repeat conformation heterogeneity in cerebellar ataxia, neuropathy, vestibular areflexia syndrome. Brain 145: 1139–1150.

Mizuguchi T, Toyota T, Miyatake S, Mitsuhashi S, Doi H, Kudo Y, Kishida H, Hayashi N, Tsuburaya RS, Kinoshita M et al. 2021. Complete sequencing of expanded SAMD12 repeats by long-read sequencing and Cas9-mediated enrichment. Brain 144: 1103–1117.

Mohren L, Erdlenbruch F, Leitão E, Kilpert F, Hönes GS, Kaya S, Schröder C, Thieme A, Sturm M, Park J et al. 2024. Advancing molecular, phenotypic and mechanistic insights of *FGF14* pathogenic expansions (SCA27B). medRxiv.

Mousavi N, Shleizer-Burko S, Yanicky R, Gymrek M. 2019. Profiling the genome-wide landscape of tandem repeat expansions. Nucleic Acids Res 47: e90.

Nemeth AH, Kwasniewska AC, Lise S, Parolin Schnekenberg R, Becker EB, Bera KD, Shanks ME, Gregory L, Buck D, Zameel Cader M et al. 2013. Next generation sequencing for molecular diagnosis of neurological disorders using ataxias as a model. Brain 136: 3106–3118.

Ngo KJ, Rexach JE, Lee H, Petty LE, Perlman S, Valera JM, Deignan JL, Mao Y, Aker M, Posey JE et al. 2020. A diagnostic ceiling for exome sequencing in cerebellar ataxia and related neurological disorders. Human mutation 41: 487–501.

Ni Y, Liu X, Simeneh ZM, Yang M, Li R. 2023. Benchmarking of Nanopore R10.4 and R9.4.1 flow cells in single-cell whole-genome amplification and whole-genome shotgun sequencing. Comput Struct Biotechnol J 21: 2352–2364.

Oberle I, Rousseau F, Heitz D, Kretz C, Devys D, Hanauer A, Boue J, Bertheas MF, Mandel JL. 1991. Instability of a 550-base pair DNA segment and abnormal methylation in fragile X syndrome. Science 252: 1097–1102.

Pais LS, Snow H, Weisburd B, Zhang S, Baxter SM, DiTroia S, O’Heir E, England E, Chao KR, Lemire G et al. 2022. seqr: A web-based analysis and collaboration tool for rare disease genomics. Human mutation 43: 698–707.

Paulson H. 2018. Repeat expansion diseases. Handbook of clinical neurology 147: 105–123.

Payne A, Holmes N, Clarke T, Munro R, Debebe BJ, Loose M. 2021. Readfish enables targeted nanopore sequencing of gigabase-sized genomes. Nature biotechnology 39: 442–450.

Pellerin D, Danzi MC, Wilke C, Renaud M, Fazal S, Dicaire MJ, Scriba CK, Ashton C, Yanick C, Beijer D et al. 2023. Deep Intronic FGF14 GAA Repeat Expansion in Late-Onset Cerebellar Ataxia. The New England journal of medicine 388: 128–141.

Porubsky D, Ebert P, Audano PA, Vollger MR, Harvey WT, Marijon P, Ebler J, Munson KM, Sorensen M, Sulovari A et al. 2021. Fully phased human genome assembly without parental data using single-cell strand sequencing and long reads. Nature biotechnology 39: 302–308.

Rafehi H, Read J, Szmulewicz DJ, Davies KC, Snell P, Fearnley LG, Scott L, Thomsen M, Gillies G, Pope K et al. 2023. An intronic GAA repeat expansion in FGF14 causes the autosomal-dominant adult-onset ataxia SCA50/ATX-FGF14. American journal of human genetics 110: 105–119.

Rafehi H, Szmulewicz DJ, Bennett MF, Sobreira NLM, Pope K, Smith KR, Gillies G, Diakumis P, Dolzhenko E, Eberle MA et al. 2019. Bioinformatics-Based Identification of Expanded Repeats: A Non-reference Intronic Pentamer Expansion in RFC1 Causes CANVAS. American journal of human genetics 105: 151–165.

Rafehi H, Szmulewicz DJ, Pope K, Wallis M, Christodoulou J, White SM, Delatycki MB, Lockhart PJ, Bahlo M. 2020. Rapid Diagnosis of Spinocerebellar Ataxia 36 in a three-Generation Family Using Short-Read Whole-Genome Sequencing Data. Mov Disord doi:10.1002/mds.28105.

Rajan-Babu IS, Dolzhenko E, Eberle MA, Friedman JM. 2024. Sequence composition changes in short tandem repeats: heterogeneity, detection, mechanisms and clinical implications. Nat Rev Genet doi:10.1038/s41576-024-00696-z.

Read JL, Davies KC, Thompson GC, Delatycki MB, Lockhart PJ. 2023. Challenges facing repeat expansion identification, characterisation, and the pathway to discovery. Emerg Top Life Sci 7: 339–348.

Rehm HL. 2017. Evolving health care through personal genomics. Nat Rev Genet 18: 259–267.

Rexach J, Lee H, Martinez-Agosto JA, Nemeth AH, Fogel BL. 2019. Clinical application of next-generation sequencing to the practice of neurology. Lancet Neurol 18: 492–503.

Richards S, Aziz N, Bale S, Bick D, Das S, Gastier-Foster J, Grody WW, Hegde M, Lyon E, Spector E et al. 2015. Standards and guidelines for the interpretation of sequence variants: a joint consensus recommendation of the American College of Medical Genetics and Genomics and the Association for Molecular Pathology. Genet Med 17: 405–424.

Rosenbohm A, Pott H, Thomsen M, Rafehi H, Kaya S, Szymczak S, Volk AE, Mueller K, Silveira I, Weishaupt JH et al. 2022. Familial Cerebellar Ataxia and Amyotrophic Lateral Sclerosis/Frontotemporal Dementia with DAB1 and C9ORF72 Repeat Expansions: An 18-Year Study. Mov Disord 37: 2427–2439.

Ruano L, Melo C, Silva MC, Coutinho P. 2014. The global epidemiology of hereditary ataxia and spastic paraplegia: a systematic review of prevalence studies. Neuroepidemiology 42: 174–183.

Rudaks LI, Yeow D, Ng K, Deveson IW, Kennerson ML, Kumar KR. 2024. An Update on the Adult-Onset Hereditary Cerebellar Ataxias: Novel Genetic Causes and New Diagnostic Approaches. Cerebellum doi:10.1007/s12311-024-01703-z.

Sadedin SP, Ellis JA, Masters SL, Oshlack A. 2018. Ximmer: a system for improving accuracy and consistency of CNV calling from exome data. Gigascience 7.

Schlotterer C, Tautz D. 1992. Slippage synthesis of simple sequence DNA. Nucleic Acids Res 20: 211–215.

Scriba CK, Stevanovski I, Chintalaphani SR, Gamaarachchi H, Ghaoui R, Ghia D, Henderson RD, Jordan N, Winkel A, Lamont PJ et al. 2023. RFC1 in an Australasian neurological disease cohort: extending the genetic heterogeneity and implications for diagnostics. Brain Commun 5: fcad208.

Stevanovski I, Chintalaphani SR, Gamaarachchi H, Ferguson JM, Pineda SS, Scriba CK, Tchan M, Fung V, Ng K, Cortese A et al. 2022. Comprehensive genetic diagnosis of tandem repeat expansion disorders with programmable targeted nanopore sequencing. Sci Adv 8: eabm5386.

Sullivan R, Chen S, Saunders CT, Yau WY, Goh YY, O’Connor E, Dominik N, Deforie VG, Morsy H, Cortese A et al. 2024. *RFC1* repeat expansion analysis from whole genome sequencing data simplifies screening and increases diagnostic rates. medRxiv doi:10.1101/2024.02.28.24303510: 2024.2002.2028.24303510.

Tankard RM, Bennett MF, Degorski P, Delatycki MB, Lockhart PJ, Bahlo M. 2018. Detecting Expansions of Tandem Repeats in Cohorts Sequenced with Short-Read Sequencing Data. American journal of human genetics 103: 858–873.

Turro E, Astle WJ, Megy K, Graf S, Greene D, Shamardina O, Allen HL, Sanchis-Juan A, Frontini M, Thys C et al. 2020. Whole-genome sequencing of patients with rare diseases in a national health system. Nature 583: 96–102.

Verkerk AJ, Pieretti M, Sutcliffe JS, Fu YH, Kuhl DP, Pizzuti A, Reiner O, Richards S, Victoria MF, Zhang FP et al. 1991. Identification of a gene (FMR-1) containing a CGG repeat coincident with a breakpoint cluster region exhibiting length variation in fragile X syndrome. Cell 65: 905–914.

Wilke C, Pellerin D, Mengel D, Traschutz A, Danzi MC, Dicaire MJ, Neumann M, Lerche H, Bender B, Houlden H et al. 2023. GAA-FGF14 ataxia (SCA27B): phenotypic profile, natural history progression and 4-aminopyridine treatment response. Brain 146: 4144–4157.

Willems T, Gymrek M, Highnam G, Genomes Project C, Mittelman D, Erlich Y. 2014. The landscape of human STR variation. Genome research 24: 1894–1904.

Zhou Y, Yuan Y, Liu Z, Zeng S, Chen Z, Shen L, Jiang H, Xia K, Tang B, Wang J. 2019. Genetic and clinical analyses of spinocerebellar ataxia type 8 in mainland China. J Neurol 266: 2979–2986.

