## Supplementary figures and images for "Bridging the gap: a prospective trial comparing programmable targeted long-read sequencing and short-read genome sequencing for genetic diagnosis of cerebellar ataxia"

### SF1

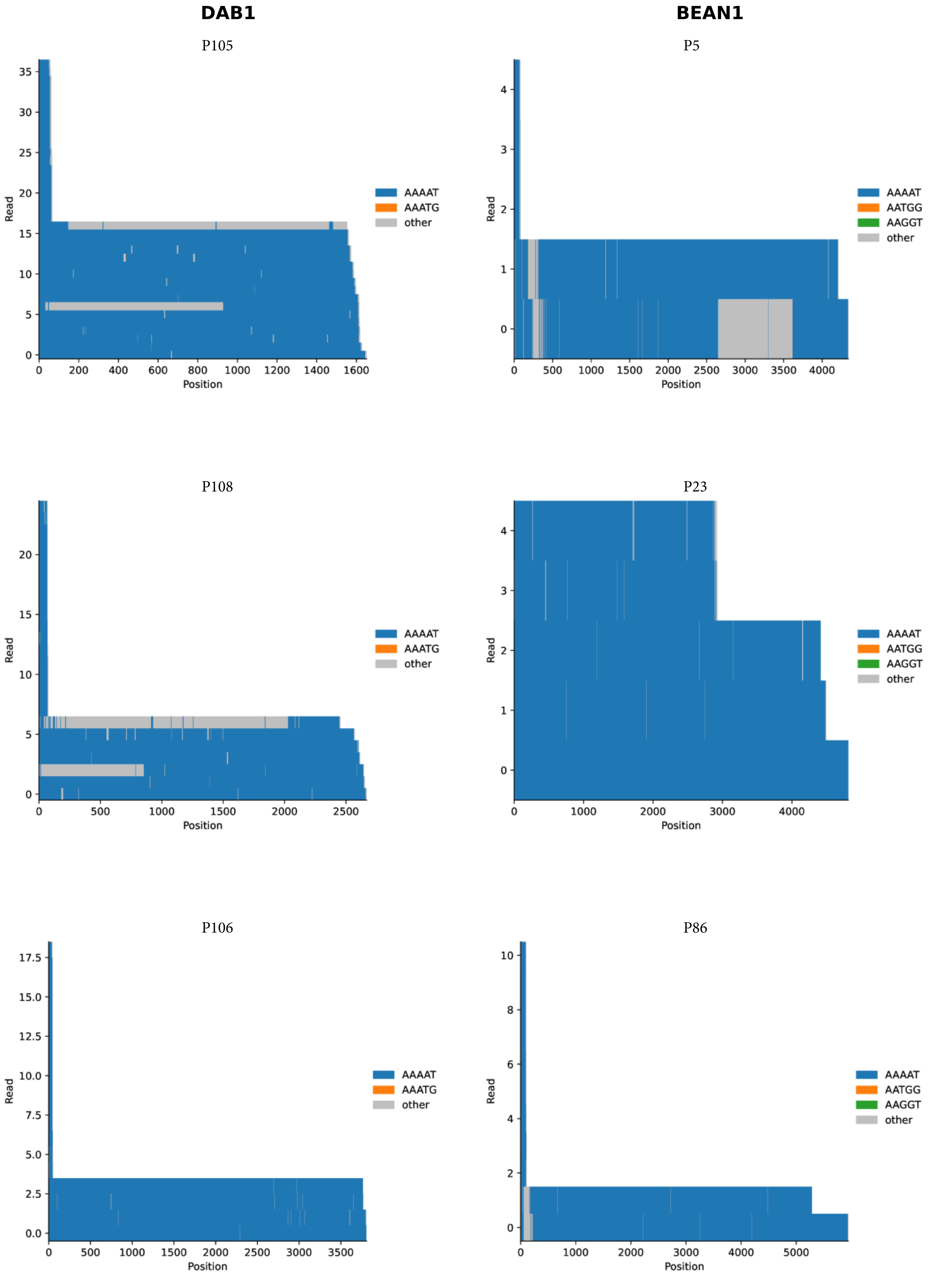

### SF2

**A** P10 – *RFC1*

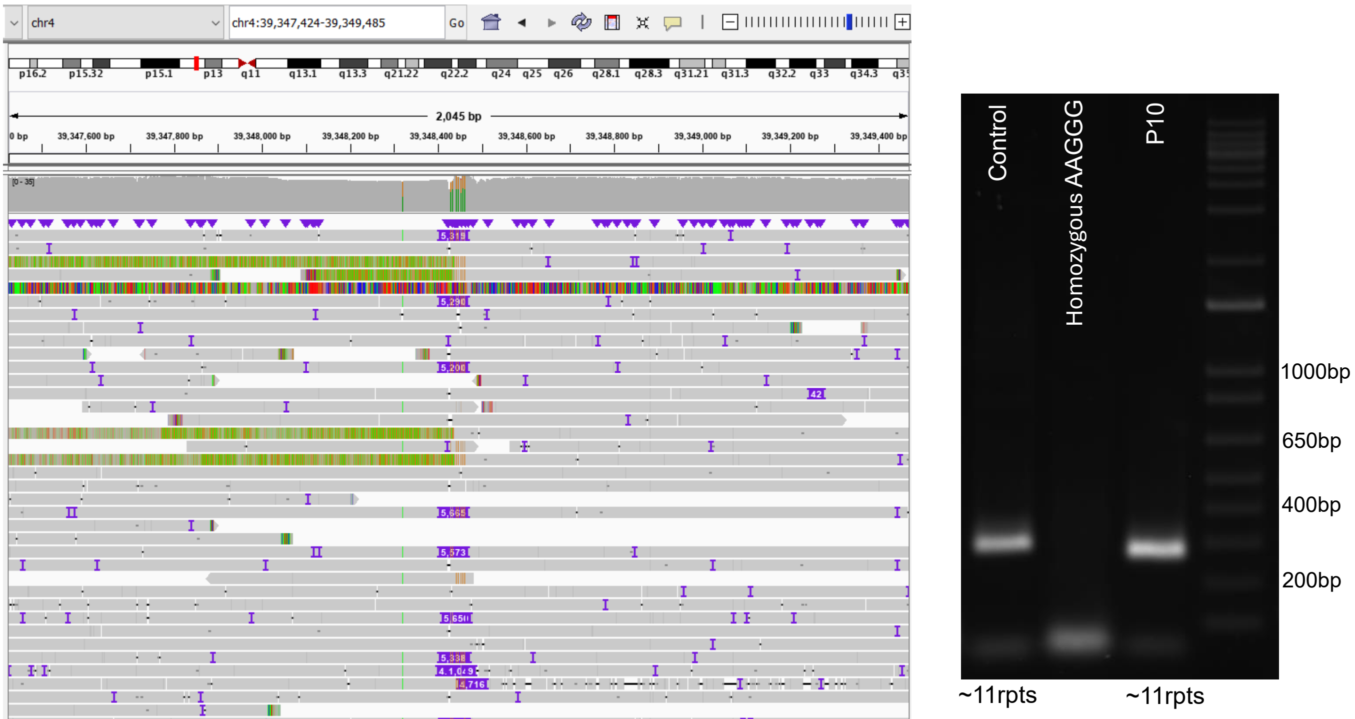


**B** P5 – *RFC1*

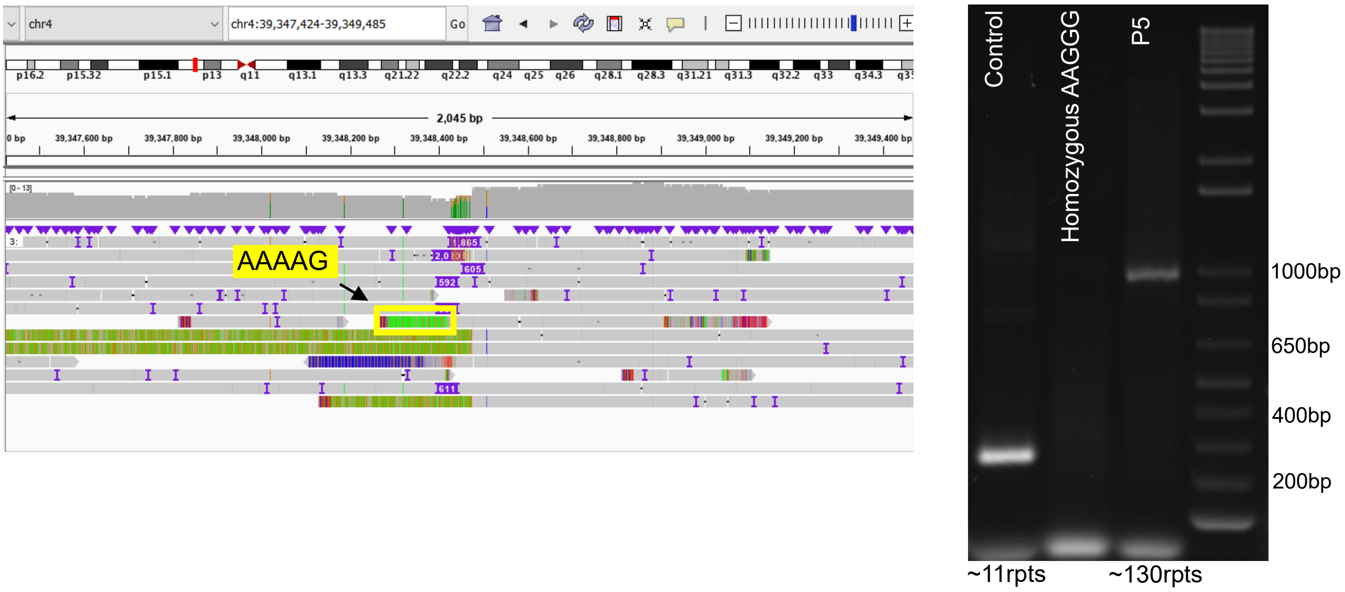


**C** P90 – *RFC1*

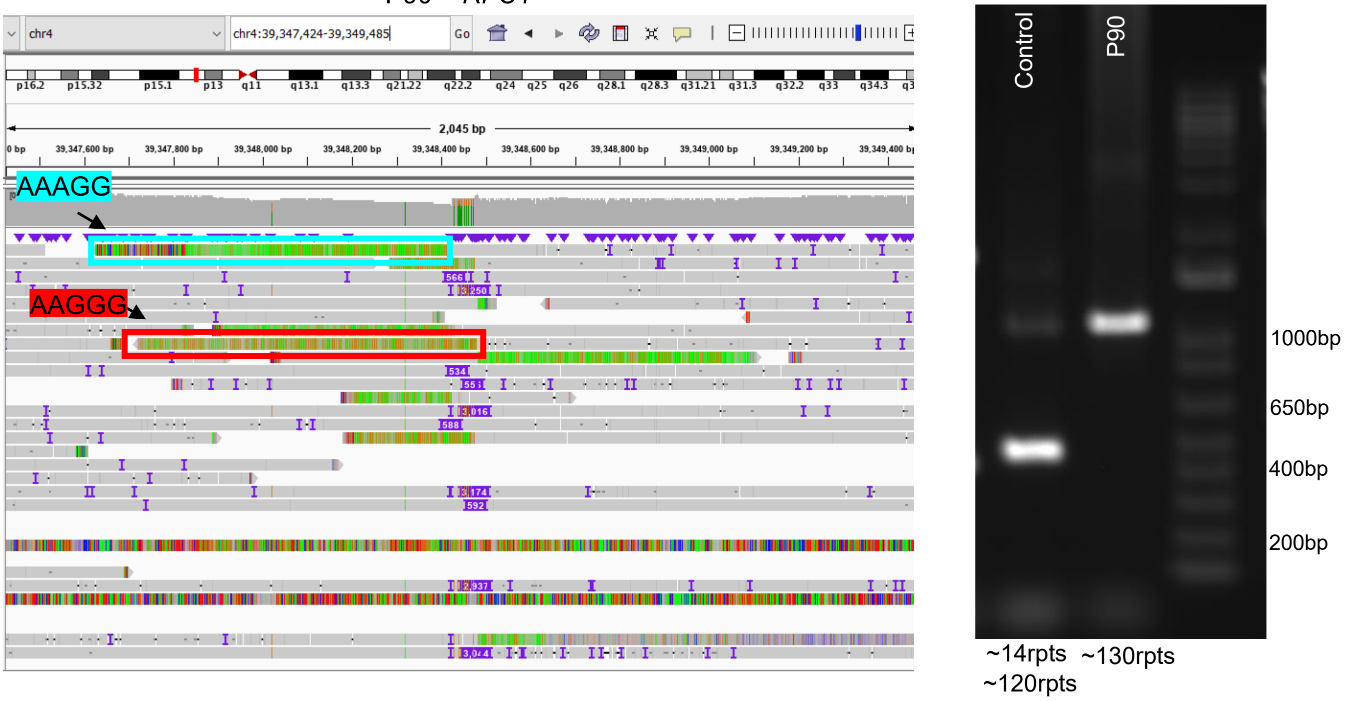

### SF3

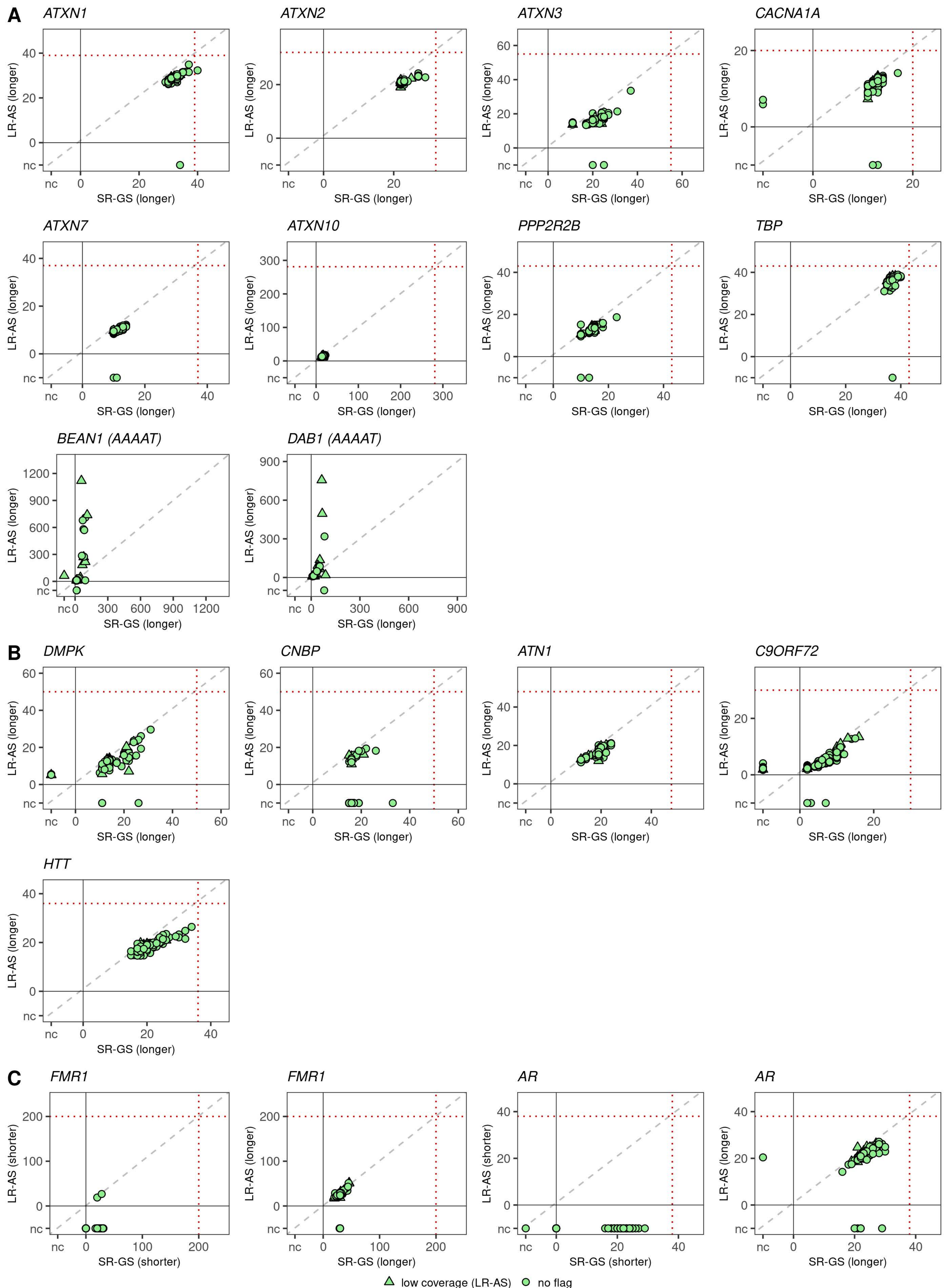

### SF4

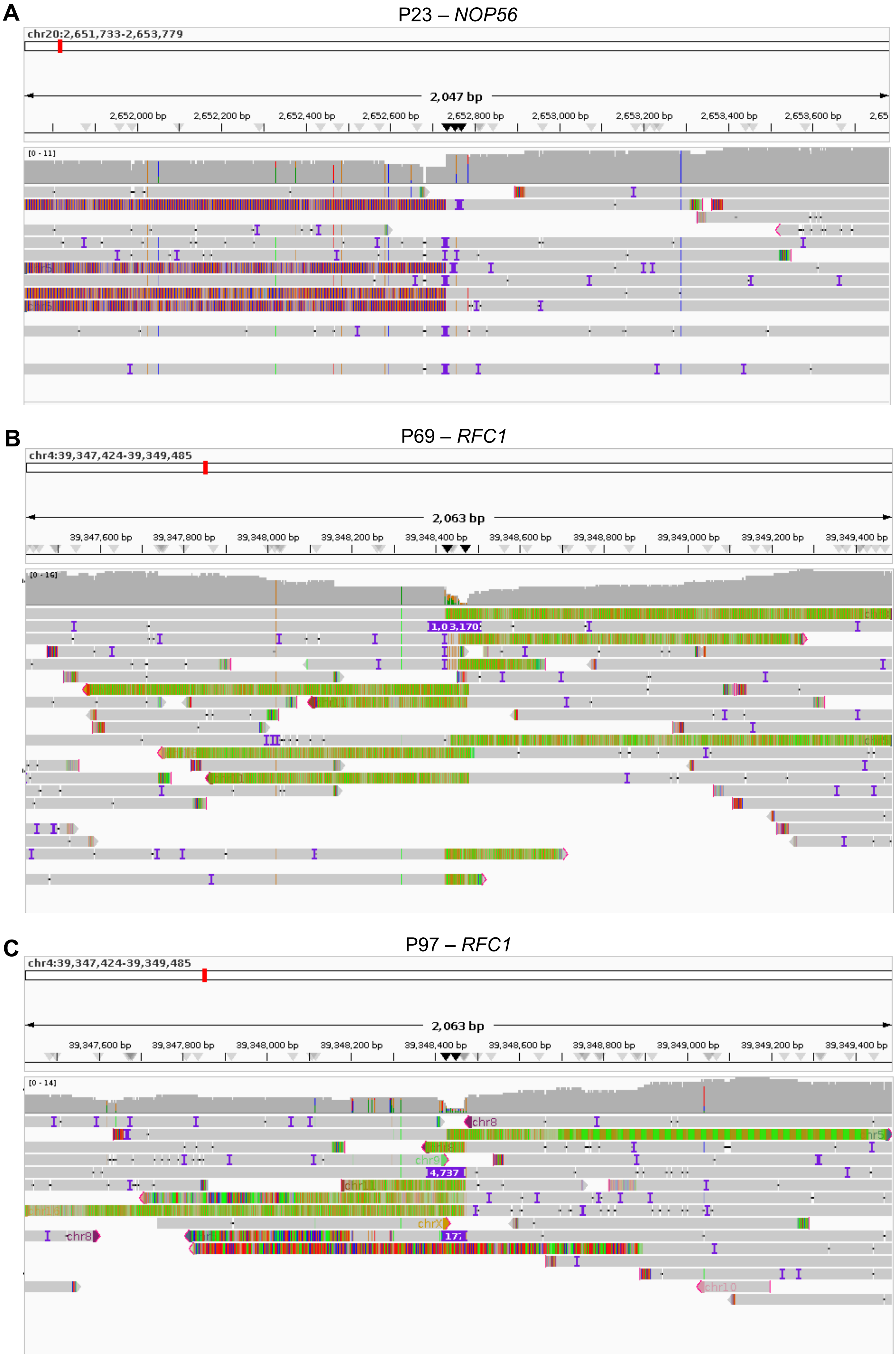

### SF5

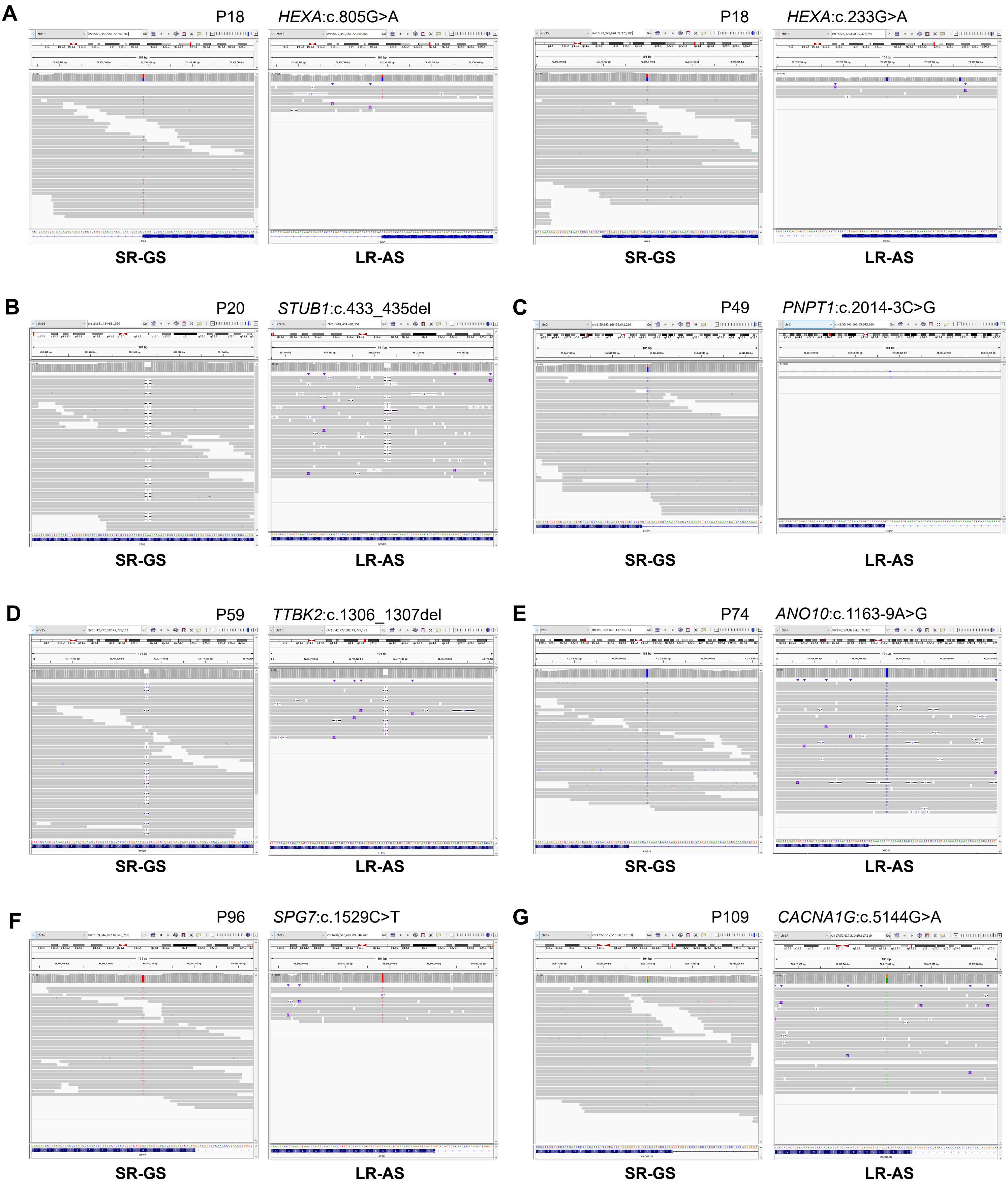
