## Supplementary material for "Bridging the gap: a prospective trial comparing programmable targeted long-read sequencing and short-read genome sequencing for genetic diagnosis of cerebellar ataxia": ST1

| **Individual ID** | **Gene** | **Disease** | **Inheritance** | **EH5 identified as expanded~** | **ONT called motif** | **ONT RE size** | **Zygosity** |
|---|---|---|---|---|---|---|---|
| P26 | *FXN* | FRDA | AR | + | (GAA)n | 14.3* | Het |
| P73 | *FXN* | FRDA | AR | + | (GAA)n | 570.9* | Het |
| P85 | *FXN* | FRDA | AR | + | $(GAA)_n$ | 629 | Het |
| P109 | *FXN* | FRDA | AR | + | $(GAA)_n$ | 160.1 | Het |
| P5 | *RFC1* | CANVAS | AR | + | $(AAGGG)_n$ | 395.4* | Het |
| P109 | *RFC1* | CANVAS | AR | + | $(AAGGG)_n$ | 1109.8* | Het |
| P14 | *RFC1* | CANVAS | AR | + | (AAAGG)n^ | 758.2* | Het |
| P47 | *RFC1* | CANVAS | AR | + | $(AAGGG)_n$ | 167.7* | Het |
| P90 | *RFC1* | CANVAS | AR | + | (AAAGG)n^ | 640.3* | Het |
| P106 | *RFC1* | CANVAS | AR | + | (AATGG)n^ | 793.3* | Het |

**Repeat expansion carrier findings for *RFC1* and *FXN***

Legend:

AR, autosomal recessive; EH5, ExpansionHunter5; +, identified; ONT, Oxford Nanopore Technologies; RE, repeat expansion; Het, heterozygous.

* less than 7 supporting reads.

^ motif incorrectly called by STRaglr. Upon manual review and orthogonal testing, motif is AAGGG.

~ without kmer filtering.
