## Supplementary material for "Bridging the gap: a prospective trial comparing programmable targeted long-read sequencing and short-read genome sequencing for genetic diagnosis of cerebellar ataxia": ST2

ST2 - Table of *FGF14* allele sizes from SR-GS, PCR and LR-AS. 1: shorter allele, 2: longer allele.

| Individual ID | SR-GS1 | PCR1 | LR-AS1 | LR-AS_reads1 | SR-GS2 | PCR2 | LR-AS2 | LR-AS_reads2 | flag |
|---|---|---|---|---|---|---|---|---|---|
| P1 | 9 | 9 | 8.7 | 3 | 87 | 169 | 168.1 | 8 | |
| P2 | 18 | 17 | 24.7 | 11 | 120 | 240 | 237.3 | 3 | |
| P3 | 134 | 182 | 154.2 | 10 | 134 | 286 | 288.5 | 10 | |
| P4 | 10 | 10 | 9.4 | 6 | 105 | 346 | 327.1 | 6 | |
| P5 | 101 | 54 | 71.9 | 3 | 101 | 308 | 328.3 | 2 | |
| P6 | 10 | 9 | 9.5 | 6 | 43 | 35 | 36.4 | 3 | |
| P7 | 9 | 9 | NA | NA | 25 | 24 | 20 | 8 | |
| P8 | 12 | 11 | NA | NA | 53 | 58 | 59.1 | 6 | |
| P9 | 12 | 11 | NA | NA | 79 | 87 | 12.7 | 7 | poor coverage in ONT |
| P10 | 10 | 9 | 9.5 | 20 | 61 | 60 | 63.5 | 20 | |
| P11 | 10 | 10 | NA | NA | 18 | 17 | 12.5 | 8 | |
| P12 | 10 | 10 | NA | NA | 10 | 10 | 9.8 | 7 | |
| P13 | 126 | 167 | 175.9 | 14 | 126 | 447 | 445.5 | 6 | |
| P14 | 10 | 9 | 9.6 | 12 | 150 | 310 | 260.9 | 9 | |
| P15 | 18 | 17 | 17.2 | 27 | 38 | 31 | 36 | 27 | |
| P16 | 10 | 10 | NA | NA | 12 | 11 | 10.6 | 50 | |
| P17 | 10 | 10 | NA | NA | 18 | 17 | 12.6 | 16 | |
| P18 | 40 | 36 | NA | NA | 40 | 39 | 38 | 7 | |
| P19 | 10 | 10 | 9.7 | 25 | 63 | 83 | 85.6 | 16 | |
| P20 | 9 | 9 | 8.7 | 23 | 41 | 39 | 40.6 | 15 | |
| P21 | 9 | 9 | 8.2 | 5 | 164 | 229 | 235.2 | 7 | |
| P22 | 114 | 71 | NA | NA | 114 | 400 | 108.3 | 5 | poor coverage in ONT |
| P23 | 18 | 17 | 15.8 | 6 | 39 | 36 | 37.6 | 9 | |
| P24 | 77 | 43 | 46.8 | 22 | 77 | 161 | 161.8 | 12 | |
| P25 | 10 | 9 | 32.5 | 11 | 118 | 315 | 311.8 | 15 | |
| P26 | 17 | 16 | NA | NA | 138 | 293 | 16.2 | 2 | poor coverage in ONT |
| P27 | 39 | 45 | NA | NA | 52 | 58 | 55.3 | 19 | |
| P28 | 17 | 16 | 15.7 | 4 | 82 | 138 | 131.1 | 4 | |
| P29 | 10 | 10 | NA | NA | 40 | 33 | NA | NA | poor coverage in ONT |
| P30 | 88 | 76 | 78.7 | 5 | 88 | 198 | 184.8 | 2 | |
| P31 | 9 | 9 | 8.8 | 3 | 36 | 38 | 39.7 | 5 | |
| P32 | 79 | 44 | 46.2 | 4 | 79 | 129 | 132.2 | 6 | |
| P33 | 10 | 9 | NA | NA | 12 | 11 | 10 | 8 | |
| P34 | 10 | 10 | NA | NA | 12 | 11 | 9.9 | 5 | |
| P35 | 9 | 9 | 8.8 | 15 | 55 | 158 | 157.3 | 16 | |
| P36 | 41 | 35 | NA | NA | 49 | 48 | 42.3 | 14 | |
| P37 | 10 | 10 | NA | NA | 17 | 16 | 12.7 | 7 | |
| P38 | 95 | 55 | 57.2 | 2 | 95 | 313 | 324.9 | 4 | |
| P39 | 10 | 9 | NA | NA | 12 | 11 | 10.6 | 40 | |
| P40 | 17 | 16 | 16.4 | 3 | 188 | 400 | 126.8 | 2 | poor coverage in ONT |
| P41 | 12 | 11 | NA | NA | 12 | 11 | 11.3 | 24 | |
| P42 | 12 | 11 | 37.7 | 5 | 126 | 253 | 252.8 | 7 | |
| P43 | 10 | 9 | NA | NA | 11 | 10 | 10.3 | 26 | |
| P44 | 10 | 9 | 9.5 | 24 | 88 | 81 | 80.9 | 22 | |
| P45 | 12 | 11 | 15 | 15 | 145 | 349 | 329.6 | 16 | |
| P46 | 17 | 16 | 16.9 | 29 | 112 | 133 | 137 | 15 | |
| P47 | 9 | 8 | NA | NA | 9 | 8 | 8.3 | 9 | |
| P48 | 17 | 16 | NA | NA | 18 | 17 | 17 | 23 | |
| P49 | 29 | 34 | NA | NA | 54 | 49 | 43.2 | 9 | |
| P50 | 10 | 9 | 9.4 | 8 | 130 | 265 | 270.6 | 4 | |
| P51 | 18 | 17 | NA | NA | 23 | 20 | 18.9 | 35 | |
| P52 | 18 | 17 | 16.9 | 5 | 117 | 281 | 277.3 | 3 | |
| P53 | 17 | 16 | 16.4 | 14 | 102 | 215 | 204.8 | 9 | |
| P54 | 10 | 9 | 9.5 | 5 | 67 | 63 | 65 | 9 | |
| P55 | 10 | 9 | NA | NA | 10 | 9 | 9.9 | 16 | |
| P56 | 10 | 9 | 9.3 | 9 | 113 | 228 | 218.7 | 6 | |
| P57 | 17 | 16 | 17.8 | 8 | 125 | 207 | 210.2 | 3 | |
| P58 | 17 | 16 | 15.9 | 5 | 45 | 39 | 41.2 | 12 | |
| P59 | 10 | 9 | NA | NA | 95 | 118 | 10 | 7 | poor coverage in ONT |
| P60 | 34 | 35 | NA | NA | 43 | 50 | 43.5 | 23 | not AAG |
| P61 | 9 | 8 | NA | NA | 9 | 8 | 8.7 | 19 | |
| P62 | 18 | 17 | 17.2 | 23 | 39 | 37 | 40 | 14 | |
| P63 | 10 | 9 | 7 | 2 | 119 | 327 | 325.4 | 3 | |
| P64 | 10 | 9 | 9.2 | 11 | 182 | 287 | 255.7 | 5 | |
| P65 | 34 | 29 | 30.3 | 17 | 62 | 54 | 55.2 | 20 | |
| P66 | 18 | 17 | 17.5 | 17 | 51 | 50 | 53 | 16 | |
| P67 | 25 | 24 | 24.6 | 13 | 186 | 267 | 265.2 | 12 | |
| P68 | 20 | 19 | 18.5 | 14 | 133 | 268 | 249.1 | 6 | |
| P69 | 10 | 9 | NA | NA | 12 | 11 | 10 | 15 | |
| P70 | 10 | 9 | NA | NA | 10 | 10 | 9.8 | 40 | |
| P71 | 40 | 38 | NA | NA | 40 | 41 | 40.7 | 13 | |
| P72 | 10 | 9 | 22.4 | 23 | 146 | 289 | 288.3 | 11 | |

| | | | | | | | | | | |
|---|---|---|---|---|---|---|---|---|---|---|
| P73 | 17 | 16 | NA | NA | | 18 | 17 | 16.4 | 12 | |
| P74 | 17 | 16 | NA | NA | | 36 | 30 | 21.4 | 33 | |
| P75 | 11 | 10 | 10.1 | | 14 | 111 | 202 | 199.5 | 7 | |
| P76 | 12 | 11 | 11.6 | | 11 | 49 | 43 | 45 | 17 | |
| P77 | 10 | 9 | NA | NA | | 12 | 11 | 10.7 | 40 | |
| P78 | 62 | 41 | 43.9 | | 15 | 62 | 305 | 316.5 | 8 | not AAG |
| P79 | 12 | 11 | 11.5 | | 8 | 82 | 113 | 114.3 | 10 | |
| P80 | 18 | 17 | 17.2 | | 18 | 45 | 40 | 42 | 16 | |
| P81 | 85 | 41 | 44.6 | | 8 | 85 | 165 | 166 | 7 | |
| P82 | 109 | 53 | 59.2 | | 17 | 109 | 230 | 231.6 | 9 | |
| P83 | 18 | 17 | 16.4 | | 16 | 41 | 34 | 35.3 | 8 | |
| P84 | 12 | 11 | NA | NA | | 18 | 17 | 14.5 | 31 | |
| P85 | 10 | 9 | 9.5 | | 9 | 57 | 82 | 85.7 | 9 | |
| P86 | 16 | 15 | NA | NA | | 17 | 16 | 15.7 | 29 | |
| P87 | 10 | 9 | NA | NA | | 17 | 16 | 13.7 | 30 | |
| P88 | 10 | 9 | NA | NA | | 10 | 9 | 9.7 | 36 | |
| P89 | 34 | 29 | 30.7 | | 14 | 118 | 146 | 148.9 | 20 | |
| P90 | 44 | 39 | 47.2 | | 11 | 109 | 315 | 310.4 | 9 | |
| P91 | 16 | 9 | NA | NA | | 18 | 15 | 12.5 | 26 | |
| P92 | 17 | 16 | 16.1 | | 36 | 102 | 135 | 135.7 | 20 | |
| P93 | 34 | 31 | NA | NA | | 39 | 32 | 33.2 | 35 | |
| P94 | 9 | 8 | NA | NA | | 9 | 9 | 8.9 | 49 | |
| P95 | 41 | 35 | NA | NA | | 47 | 39 | 40 | 41 | |
| P96 | 12 | 11 | 11.6 | | 11 | 44 | 43 | 45.5 | 13 | |
| P97 | 10 | 9 | NA | NA | | 17 | 16 | 14.5 | 18 | |
| P98 | 17 | 16 | 16.6 | | 17 | 57 | 120 | 120.5 | 19 | |
| P99 | 10 | 9 | 14.9 | | 25 | 61 | 88 | 149 | 7 | not AAG |
| P100 | 90 | 54 | 76.6 | | 27 | 90 | 374 | 378 | 9 | |
| P101 | 43 | 35 | 47.6 | | 29 | 151 | 317 | 316.4 | 20 | |
| P102 | 36 | 19 | 19.1 | | 20 | 44 | 36 | 37.9 | 21 | |
| P103 | 12 | 11 | 11 | | 13 | 134 | 159 | 153.9 | 12 | |
| P104 | 11 | 10 | 10.6 | | 15 | 68 | 54 | 56.3 | 25 | |
| P105 | 10 | 9 | 15.8 | | 14 | 100 | 208 | 205.1 | 15 | |
| P106 | 18 | 17 | 26.6 | | 14 | 148 | 310 | 316.4 | 8 | |
| P107 | 101 | 52 | 62.2 | | 19 | 101 | 322 | 325.3 | 19 | |
| P108 | 75 | 41 | 43.5 | | 17 | 75 | 134 | 133.7 | 14 | |
| P109 | 10 | 9 | 9.5 | | 7 | 53 | 44 | 44.9 | 16 | |
| P110 | 10 | 9 | NA | NA | | 12 | 11 | 10.4 | 39 | |
