## Supplementary material for "Bridging the gap: a prospective trial comparing programmable targeted long-read sequencing and short-read genome sequencing for genetic diagnosis of cerebellar ataxia": ST3

| Individual ID | Sex | Age at testing | Age at onset | FHx of CA | Gene | Disease | Inheritance | Variant motif | EH5 RE size | ONT RE size | PCR RE size | Zygosity | Clinical presentation |
|---|---|---|---|---|---|---|---|---|---|---|---|---|---|
| P2 | M | 9th Decade | 6th Decade | - | *FGF14* | SCA27B | AD | $(GAA)_n$ | 120 | 237.3* | 240 | Het | CABV |
| P21 | F | 8th Decade | 8th Decade | - | *FGF14* | SCA27B | AD | $(GAA)_n$ | 164 | 235.2 | 229 | Het | CA |
| P30 | M | 7th Decade | 6th Decade | - | *FGF14* | SCA27B | AD | $(GAA)_n$ | 88 | 184.8* | 198 | Het | CA |
| P53 | F | 9th Decade | 8th Decade | - | *FGF14* | SCA27B | AD | $(GAA)_n$ | 102 | 204.8 | 215 | Het | CABV |
| P56 | M | 9th Decade | 8th Decade | - | *FGF14* | SCA27B | AD | $(GAA)_n$ | 113 | 218.7* | 228 | Het | CABV |
| P57 | F | 4th Decade | 4th Decade | - | *FGF14* | SCA27B | AD | $(GAA)_n$ | 125 | 210.2* | 207 | Het | CA |
| P75 | F | 8th Decade | ND | - | *FGF14* | SCA27B | AD | $(GAA)_n$ | 111 | 199.5 | 202 | Het | CA |
| P82 | F | 9th Decade | 8th Decade | - | *FGF14* | SCA27B | AD | $(GAA)_n$ | 109 | 231.6 | 230 | Het | EA |
| P105 | M | 8th Decade | 8th Decade | - | *FGF14* | SCA27B | AD | $(GAA)_n$ | 100 | 205.1 | 208 | Het | deafness |

**Repeat expansion variants of uncertain signficance identified.**

Legend:
M, male; F, female: ND, no data available; FHx, family history; CA, cerebellar ataxia; -, absent; AD, autosomal dominant; EH5, ExpansionHunter5; RE, repeat expansion; ONT, Oxford Nanopore Technologies
Het, heterozygous; CABV, cerebellar ataxia and bilateral vestibulopathy; EA, episodic ataxia
* less than 7 supporting reads.
