## Supplementary material for "Bridging the gap: a prospective trial comparing programmable targeted long-read sequencing and short-read genome sequencing for genetic diagnosis of cerebellar ataxia": ST4

| Individual ID | Sex | testing | Age at onset | FHx of CA | Gene | Disease | Inheritance | Genomic variant (hg38) | HGVSc | HGVSp | Zygosity | Consequence | Classification | Clinical presentation |
|---|---|---|---|---|---|---|---|---|---|---|---|---|---|---|
| P17 | M | 5th Decade | 4th Decade | + | *CACNA1A* | disorder | AD | chr19:13259654C>T | c.4298G>A | p.(Arg1433Gln) | Het | missense | VUS | microduplication) |
| P98 | M | 6th Decade | 5th Decade | + | *CACNA1A* | disorder | AD | chr19:13455156A>G | c.350T>C | p.(Leu117Pro) | Het | missense | VUS | CA |
| P47 | F | 7th Decade | 7th Decade | - | *CACNA1G* | SCA42 | AD | chr17:50604166G>A | c.4181G>A | p.(Arg1394Gln) | Het | missense | VUS | CA |
| P58 | M | 6th Decade | 5th Decade | - | *KCND3* | SCA19/22 | AD | >A | c.1882del | ) | Het | frameshift | VUS | CA and tremor |
| P89 | M | 8th Decade | 7th Decade | - | *KCND3* | SCA19/22 | AD | chr1:111982684C>T | c.43G>A | p.(Ala15Thr) | Het | missense | VUS | CA and upper limb hyperreflexia |
| P11 | F | 9th Decade | 7th Decade | + | *NPTX1* | SCA50 | AD | chr17:80475584G>C | c.579C>G | p.(Asn193Lys) | Het | missense | VUS | CA and Meniere's disease |
| P27 | M | 8th Decade | 8th Decade | + | *NPTX1* | SCA50 | AD | chr17:80475584G>C | c.579C>G | p.(Asn193Lys) | Het | missense | VUS | CA and right cataract |
| P51 | F | 7th Decade | 6th Decade | - | *PRKCG* | SCA14 | AD | chr19:53891737T>C | c.593T>C | p.(Leu198Pro) | Het | missense | VUS | CA |
| P55 | F | 7th Decade | ND | - | *STUB1* | SCA48 | AD | chr16:681475GGAC>G | c.400_402del | p.(Asp134del) | Het | inframe deletion | VUS | CA and dementia |

**Single nucleotide and small insertion or deletion variants of uncertain significance.**

Legend:

M, male; F, female; ND, no data available; FHx, family history; CA, cerebellar ataxia; -, absent; +, present; AD, autosomal dominant; Het, heterozygous; VUS, variant of uncertain significance; ID, intellectual disability.

Transcripts: *CACNA1A* , NM_001127222.2; *CACNA1G* , NM_018896.5; *FAT2* , NM_001447.3; *KCND3* , NM_001378969.1; *NPTX1* , NM_002522.4; *PRKCG* , NM_002739.5; *STUB1* , NM_005861.4.
