## Supplementary material for "Bridging the gap: a prospective trial comparing programmable targeted long-read sequencing and short-read genome sequencing for genetic diagnosis of cerebellar ataxia": ST6

| Individual ID | Gene | Disease | Diagnostic RE sizing* (Allele 1) | ONT RE size (Allele 1) | ONT RE size supporting reads | Diagnostic RE sizing* (Allele 2) | ONT RE size (Allele 2) | ONT RE size supporting reads |
|---|---|---|---|---|---|---|---|---|
| C1 | *ATXN1* | SCA1 | 52-54 | 48.4 | 14 | 36-38 | 28.6 | 12 |
| C2 | *ATXN2* | SCA2 | 42 | 43.3 | 8 | 21 | 18.5 | 11 |
| C3 | *ATXN3* | SCA3 | 66 | 61 | 14 | 18 | 13.7 | 15 |
| C4 | *CACNA1A* | SCA6 | 23 | 14.4# | 14 | 12 | No call | No call |
| C5 | *ATXN7* | SCA7 | 39 | 37 | 2 | 13 | 7.7 | 5 |

**Long-read adaptive sequencing validation of diagnostic testing for SCA1,2,3,6,7**

RE, repeat expansion; ONT Oxford Nanopore Technologies.

*clinical diagnostic PCR testing, +/- 2 repeats.

### STRaglr only sized a single allele and did not call the allele as expanded, manual inspection in IGV showed 4 reads with CAG22-23 and 10 reads with CAG12.
